# Personalized survival probabilities for SARS-CoV-2 positive patients by explainable machine learning

**DOI:** 10.1101/2021.10.28.21265598

**Authors:** Adrian G. Zucco, Rudi Agius, Rebecka Svanberg, Kasper S. Moestrup, Ramtin Z. Marandi, Cameron Ross MacPherson, Jens Lundgren, Sisse R. Ostrowski, Carsten U. Niemann

## Abstract

Interpretable risk assessment of SARS-CoV-2 positive patients can aid clinicians to implement precision medicine. Here we trained a machine learning model to predict mortality within 12 weeks of a first positive SARS-CoV-2 test. By leveraging data on 33,928 confirmed SARS-CoV-2 cases in eastern Denmark, we considered 2,723 variables extracted from electronic health records (EHR) including demographics, diagnoses, medications, laboratory test results and vital parameters. A discrete-time framework for survival modelling enabled us to predict personalized survival curves and explain individual risk factors. Performances of weighted concordance index 0.95 and precision-recall area under the curve 0.71 were measured on the test set. Age, sex, number of medications, previous hospitalizations and lymphocyte counts were identified as top mortality risk factors. Our explainable survival model developed on EHR data also revealed temporal dynamics of the 22 selected risk factors. Upon further validation, this model may allow direct reporting of personalized survival probabilities in routine care.

## INTRODUCTION

Coronavirus disease 2019 (COVID-19) caused by infection with Severe acute respiratory syndrome coronavirus 2 (SARS-CoV-2) has by October 2021 claimed almost 5 million lives since its outbreak in late 2019^1^. Infected individuals present a variety of symptoms, ranging from asymptomatic to life-threatening diseases^2^. Although the majority of cases experience mild to moderate disease approximately 15% of confirmed SARS-CoV-2 positive cases are estimated to develop severe disease^3^. Progression to severe disease seems to occur within 1-2 weeks from symptom onset, and is characterized by clinical signs of pneumonia with dyspnea, increased respiratory rate, and decreased blood oxygen saturation requiring supplemental oxygen^3–7^. Development of critical illness is driven by systemic inflammation, leading to acute respiratory distress syndrome (ARDS), respiratory failure, septic shock, multi-organ failure, and/or disseminated coagulopathy^4,5,8^. The majority of these patients require mechanical ventilation, and mortality for patients admitted to an Intensive Care Unit (ICU) is reported to be 32-50%^3,8–10^. Despite the current vaccination program, both people already vaccinated and patients not being vaccinated continue to develop critical COVID-19 disease^11^. Thus, the pandemic still poses a great burden on health care systems worldwide, locally approaching the limit of capacity due to high patient burden and challenging clinical management.

Several factors associated with increased risk of severe disease course have been established including old age, male gender, and lifestyle factors such as smoking and obesity^12,13^. Comorbidities including hypertension, type 2 diabetes, renal disease, as well as pre-existing conditions of immune dysfunction and cancer, are also associated with a higher risk of severe disease and COVID-19 related death^12,14–16^. Among hospitalized patients, risk factors for severe disease or death include low lymphocyte counts, elevated inflammatory markers and elevated kidney and liver parameters indicating organ dysfunction^6^. However, many of these factors likely reflect an ongoing progression of COVID-19. Thus, identification of high-risk patients at or prior to hospital admission is warranted to facilitate personalized interventions.

Multiple COVID-19 prognostic models have been built on reduced sets of predictive features from demographics, patient history, physical examination, and laboratory results^17^ processed by traditional statistical frameworks or machine learning (ML) algorithms. A systematic review of 50 prognostic models has concluded that overall such models have been poorly reported and are at a high risk of bias^18^. While great efforts have been put into providing prognostic models based on data collected from health systems, traditional modelling approaches solely based on domain knowledge may fail. This represents a risk of missing novel markers and insights about the disease that could come from data-driven models in a hypothesis-free manner^19^, which have been reported to outperform models based on curated variables from domain experts^20^.

Furthermore, ML models facilitate clinical insights^21^ when coupled with methods for model explainability such as SHapley Additive exPlanations (SHAP) values^22^. Model explainability has been developed mainly in the context of regression and binary classification, but in clinical research where censored observations are common, explainable time-to-event modelling is required to avoid selection bias^23,24^. Multiple ML algorithms have been developed for time-to-event modelling, either by building on top of existing models such as Cox proportional hazards or by defining new loss functions that model time as continuous^25^. Here we used an alternative approach that considered time in discrete intervals and performed binary classification at such time intervals^26^. This allowed us to implement gradient boosting decision trees for binary classification to predict personalized survival probabilities^27^ and allow explainability at the individual patient level using SHAP values^22^ including temporal dynamics of risk factors over the course of the disease. This approach not only allows to predict personalized survival probabilities and risk factors for SARS-CoV-2 positive patients but also provides a framework for precision medicine that can be applied to other diseases based on routine electronic health records.

## RESULTS

### Patient cohort

Based on centralized EHR and SARS-CoV-2 test results from test centers in eastern Denmark, we identified 33,938 patients who had at least one SARS-CoV-2 RT-PCR positive test from 963,265 individuals who had a test performed between 17th of March 2020 and 2^nd^ of March 2021 (Fig. 1). In this cohort, 5,077 patients were hospitalized, of whom 502 were admitted to the ICU (Supplementary Fig. 1). Overall, 1,803 (5.34%) deaths occurred among all individuals with a positive SARS-CoV-2 RT-PCR test, of whom 141 died later than 12 weeks from the first positive test (FPT) hence considered as alive for this analysis. Right-censoring was only observed for patients tested after the 8^th^ of December 2020 with less than 12 weeks of follow-up available while deaths that occurred the same day of FPT were not considered for training. For the initial model, demographics, laboratory test results, hospitalizations, vital parameters, diagnoses, medicines (ordered and administered) and summary features were included. Feature encoding resulted in 2,723 features (Supplementary Table 2) which after feature selection were reduced to 23 features. A summary of the cohort based on the final feature set can be found in Table 1. This cohort represents an updated subset of individuals residing in Denmark characterized in a previous publication^28^.

**Figure 1.**
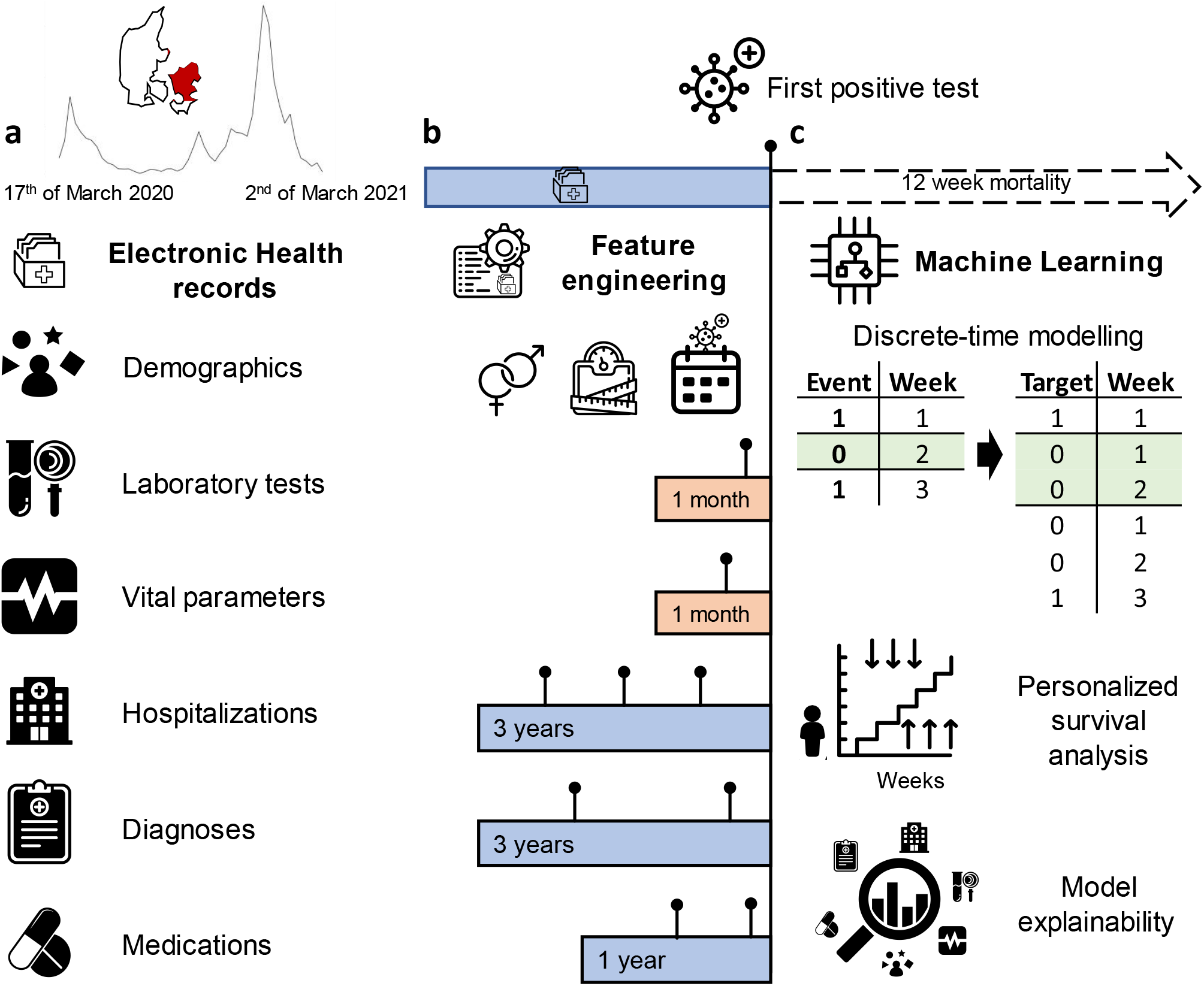
Overview of the data sources, feature engineering and modelling approach for predicting 12-week mortality in SARS-CoV-2 positive patients. **a**, Electronic Health Records (EHR) of 33,938 patients from 17^th^ of March 2020 to 2^nd^ of March 2021 (incidence curve) in eastern Denmark (geographical region visualized in red) were used to predict 12-week mortality from the first positive SARS-CoV-2 test (FPT). **b**, Features were engineered as the last value observed prior to FPT within the last month for vitals and laboratory values. To encode hospital admissions, medications and diagnoses, the count of occurrences within three or one year(s) prior to FPT was used. **c**, Machine learning algorithms were trained for survival modelling using a discrete-time approach. Time-to-event data were transformed longitudinally into patient-weeks up to the loss of follow-up (0) or death (1). With the augmented data, binary classification was performed by gradient boosting decision trees to predict personalized survival distributions for each patient and provide explanations of individual risk factors using SHAP values.

**Table 1.** Summary statistics of the cohort based on the final feature set. Values up to the day of the first positive SARS-CoV-2 test used for training and prediction were considered. Continuous variables were summarized by the median and interquartile ranges (Q1, Q3). Diagnoses and medicines with their ICD-10 and ATC codes in parentheses respectively were summarized as the number of patients with at least one code assigned. Only body mass index and absolute lymphocyte counts reported missing values for 17,823 and 32,803 patients respectively. Patients that had a positive test from the 8^th^ of December 2020 (12-weeks before data generation) and did not die before the 2^nd^ of March 2021 were censored.

|  | Level | Overall | Censored | Died | Survived |
| --- | --- | --- | --- | --- | --- |
| n |  | 33938 | 14907 | 1662 | 17369 |
| Age, median [Q1,Q3] |  | 49.0 [33.0,64.0] | 50.0 [34.0,66.0] | 83.0 [75.0,89.0] | 45.0 [31.0,59.0] |
| Sex, n (%) | Female | 19581 (57.7) | 8800 (59.0) | 787 (47.4) | 9994 (57.5) |
| Number of ordered medicines, median [Q1,Q3] |  | 0.0 [0.0,4.0] | 0.0 [0.0,4.0] | 16.0 [4.0,27.0] | 0.0 [0.0,2.0] |
| n (%) | 0 | 20207 (59.5) | 8854 (59.4) | 168 (10.1) | 11185 (64.4) |
|  | >= 1 | 13731 (40.5) | 6053 (40.6) | 1494 (89.9) | 6184 (35.6) |
| Number of diagnoses, median [Q1,Q3] |  | 4.0 [2.0,8.0] | 4.0 [2.0,8.0] | 11.0 [6.0,16.0] | 4.0 [2.0,7.0] |
| n (%) | 0 | 5596 (16.5) | 2277 (15.3) | 54 (3.2) | 3265 (18.8) |
|  | >= 1 | 28342 (83.5) | 12630 (84.7) | 1608 (96.8) | 14104 (81.2) |
| Admitted at the time of first positive test, n (%) |  | 2485 (7.3) | 927 (6.2) | 534 (32.1) | 1024 (5.9) |
| Previous admissions in the last 3 years, median [Q1,Q3] |  | 0.0 [0.0,1.0] | 0.0 [0.0,1.0] | 1.0 [0.0,3.0] | 0.0 [0.0,0.0] |
| Cumulative days in hospital within the last 3 years, median [Q1,Q3] |  | 0.0 [0.0,1.0] | 0.0 [0.0,1.0] | 7.0 [0.0,19.0] | 0.0 [0.0,0.0] |
| Pandemic week, median [Q1,Q3] |  | 39.0 [19.0,42.0] | 42.0 [41.0,44.0] | 40.0 [7.0,43.0] | 26.0 [6.0,35.0] |
| Body Mass Index, median [Q1,Q3] |  | 25.7 [22.6,29.7] | 25.6 [22.5,29.8] | 24.3 [21.3,28.0] | 26.0 [23.0,29.9] |
| Absolute Lymphocyte count (LYM), laboratory, last value, median [Q1,Q3] |  | 1.1 [0.7,1.6] | 1.1 [0.8,1.7] | 0.9 [0.6,1.3] | 1.2 [0.8,1.6] |
| Laxatives (A06AD), Ordered Medicine, count, n (%) | 0 | 31131 (91.7) | 13639 (91.5) | 919 (55.3) | 16573 (95.4) |
|  | >= 1 | 2807 (8.3) | 1268 (8.5) | 743 (44.7) | 796 (4.6) |
| Paracetamol (N02BE), Ordered Medicine, count, n (%) | 0 | 26453 (77.9) | 11430 (76.7) | 527 (31.7) | 14496 (83.5) |
|  | >= 1 | 7485 (22.1) | 3477 (23.3) | 1135 (68.3) | 2873 (16.5) |
| Loop diuretics (C03CA), Ordered Medicine, count, n (%) | 0 | 31403 (92.5) | 13753 (92.3) | 965 (58.1) | 16685 (96.1) |
|  | >= 1 | 2535 (7.5) | 1154 (7.7) | 697 (41.9) | 684 (3.9) |
| Opioid anesthetics (N01AH), Ordered Medicine, count, n (%) | 0 | 30920 (91.1) | 13470 (90.4) | 1314 (79.1) | 16136 (92.9) |
|  | >= 1 | 3018 (8.9) | 1437 (9.6) | 348 (20.9) | 1233 (7.1) |
| Vitamin B-complex (A11EA), Ordered Medicine, count, n (%) | 0 | 33216 (97.9) | 14515 (97.4) | 1497 (90.1) | 17204 (99.1) |
|  | >= 1 | 722 (2.1) | 392 (2.6) | 165 (9.9) | 165 (0.9) |
| Alzheimer's disease (G30), Diagnose, count, n (%) | 0 | 33432 (98.5) | 14655 (98.3) | 1529 (92.0) | 17248 (99.3) |
|  | >= 1 | 506 (1.5) | 252 (1.7) | 133 (8.0) | 121 (0.7) |
| Encounter for medical observation (Z03), Diagnose, count, n (%) | 0 | 24170 (71.2) | 10568 (70.9) | 788 (47.4) | 12814 (73.8) |
|  | >= 1 | 9768 (28.8) | 4339 (29.1) | 874 (52.6) | 4555 (26.2) |
| Encounter for other special examination (Z01), Diagnose, count, n (%) | 0 | 18284 (53.9) | 7485 (50.2) | 637 (38.3) | 10162 (58.5) |
|  | >= 1 | 15654 (46.1) | 7422 (49.8) | 1025 (61.7) | 7207 (41.5) |
| Essential hypertension (I10), Diagnose, count, n (%) | 0 | 31501 (92.8) | 13783 (92.5) | 1256 (75.6) | 16462 (94.8) |
|  | >= 1 | 2437 (7.2) | 1124 (7.5) | 406 (24.4) | 907 (5.2) |
| Loop diuretics (C03CA), Administered medicine, count, n (%) | 0 | 32384 (95.4) | 14135 (94.8) | 1177 (70.8) | 17072 (98.3) |
|  | >= 1 | 1554 (4.6) | 772 (5.2) | 485 (29.2) | 297 (1.7) |
| Calcium + vitamin D (A12AX), Administered medicine, count, n (%) | 0 | 32530 (95.9) | 14200 (95.3) | 1267 (76.2) | 17063 (98.2) |
|  | >= 1 | 1408 (4.1) | 707 (4.7) | 395 (23.8) | 306 (1.8) |

### Survival modelling with machine learning achieves high discriminative performance

To predict the risk of death within 12 weeks from FPT, we trained gradient boosting decision trees considering time as discrete in a time-to-event framework. Performance was measured on 20% of the data (test set) unblinded only for performance assessment. The weighted concordance index (C-index) for predicting risk of death for all 12 weeks with 95% confidence intervals (CI) was 0.946 (0.941-0.950). Binary metrics were calculated for each predicted week by excluding censored individuals (Fig. 2). At week 12, the precision-recall area under the curve (PR-AUC) and Mathew correlation coefficient (MCC) with 95% CI were 0.686 (0.651-0.720) and 0.580 (0.562-0.597) respectively. The sensitivity was 99.3% and the specificity was 86.4%. The performance for subgroups of patients displayed some differences. In patients tested outside the hospital (Fig 2b), the C-index was 0.955 (0.950-0.960), the PR-AUC and MCC were 0.675 (0.632-0.719) and 0.585 (0.562-0.605) respectively. 98.9% sensitivity and 89.9% specificity were measured in this group. For patients previously admitted to the hospital at the time of test (Fig. 2c), the C-Index was 0.809 (0.787-0.829), the PR-AUC and MCC were 0.705 (0.640-0.760) and 0.357 (0.325-0.387) respectively. The sensitivity was 100% and the specificity 31.0% indicating a higher number of false positives when using a 0.5 probability threshold for this group (Supplementary Table 1).

**Figure 2.**
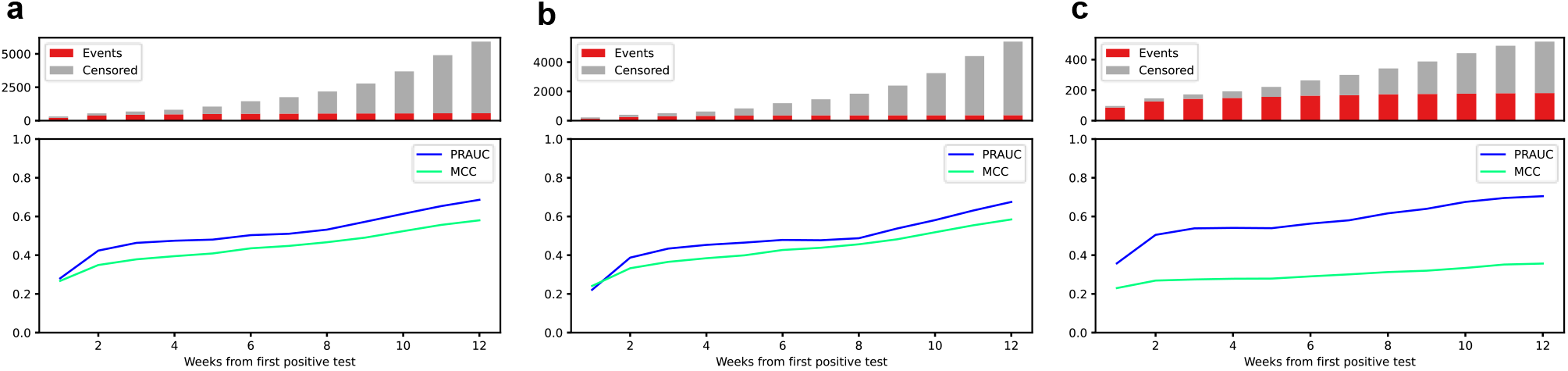
Binary performance metrics for 12 weeks mortality prediction. Precision-recall area under the curve (PR-AUC) and Mathews correlation coefficient (MCC) were calculated for each predicted week only considering non-censored patients in the test set. The lower panel of each plot depicts the mean values of PR-AUC and MCC at each week based on all patients (**a**), patients not admitted to the hospital at the time of first positive test (**b**) and patients who were admitted at the time of first positive test (**c**). The upper panels of each subfigure contain bar plots showing the number of patients who died (red) during the given week while patients censored due to lack of follow-up (grey) were omitted for the performance metrics.

### Predicted individual survival distributions represent patients’ heterogeneity

Individual survival distributions were predicted for patients in the test set. The median of the predicted cumulative death probabilities by survival status reflected the discriminative performance of the individual survival predictions (Fig. 3a). Deceased patients exhibited a risk of mortality that increased for the first month after FPT. Patients who died 2 months after the FPT exhibited a higher instant risk of death at these later periods than those patients who died earlier (Fig 3b). Our survival modelling approach is also able to approximate the time of death within the 12-week time window, as highlighted by the predicted discrete (Fig 3c) and cumulative death probabilities (Fig 3d) for three individual patients. Early death was observed as a steep increase in death probability in the first weeks while late death was observed as a gradual increase in cumulative death probability (Fig 3c). Our modelling approach also considered censored patients for which death probabilities were predicted for all periods even after censoring (Fig 3c-d)

**Figure 3.**
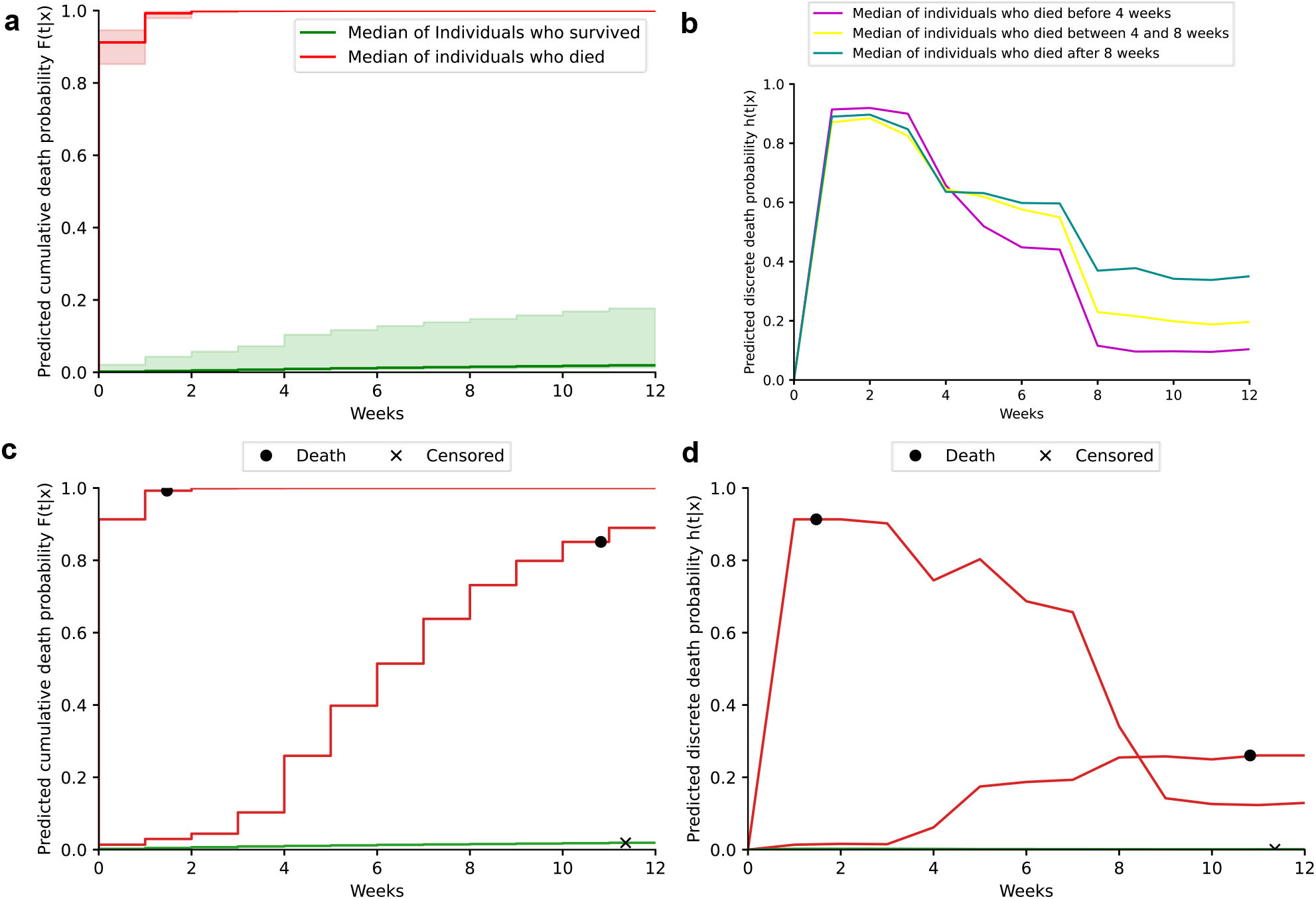
Predicted individual discrete and cumulative death probabilities. Weekly discrete and cumulative probabilities of death were predicted for all patients in the test set using data prior to their first positive test. Individual probabilities were summarized by the median, 80 and 20 percentiles for patients who died (red) or survived (green) (**a**). Predicted cumulative death probabilities were summarized by the median (**b**) for patients who died before 4 weeks (pink), between 4 and 8 weeks (yellow) and after 8 weeks (blue). Individual examples of predicted cumulative (**c**) and discrete (**d**) death probabilities for three patients are depicted indicating the time of death (black dot) or censoring (x).

### Local and global model explainability reveal temporal dynamics of mortality risk factors

Feature selection for the final model was data-driven using 5-fold cross-validation on the training set. From the original set of 2,723 features generated from routine EHR data (Supplementary Table 2), 22 features were selected. This selection was based on feature importance filtering represented by the mean of absolute SHAP values. Among top features, basic characteristics such as age, BMI, and sex, as well as clinical factors such as the number of different prescribed medications and diagnosis codes were represented (Fig. 4a). Moreover, hospitalization at the time of FPT was identified to impact the risk of death. This is further emphasized by the different performances of the model when restricted to this sub-cohort (Supplementary Table 1). We also identified the week during the pandemic in which the FPT was taken as having an impact on the risk of death. Furthermore, the risk of death was higher within the first four weeks after FPT as encoded by the week from the prediction feature. The model also allowed us to explore the temporal dynamics of individual risk factors across the predictive 12-weeks window (Fig 4b). Features such as age, ordered loop diuretics, and admission at the time of FPT had a higher impact on the risk of dying early, while BMI, diagnosis of Alzheimer’s disease, and ordered B-vitamin contributed more to late risk. Thus, identification of such time dependency for features at the individual patient level further reveals different risk factors acting on different time-horizons for the predicted risk of individual patients (Fig 4c-d).

**Figure 4.**
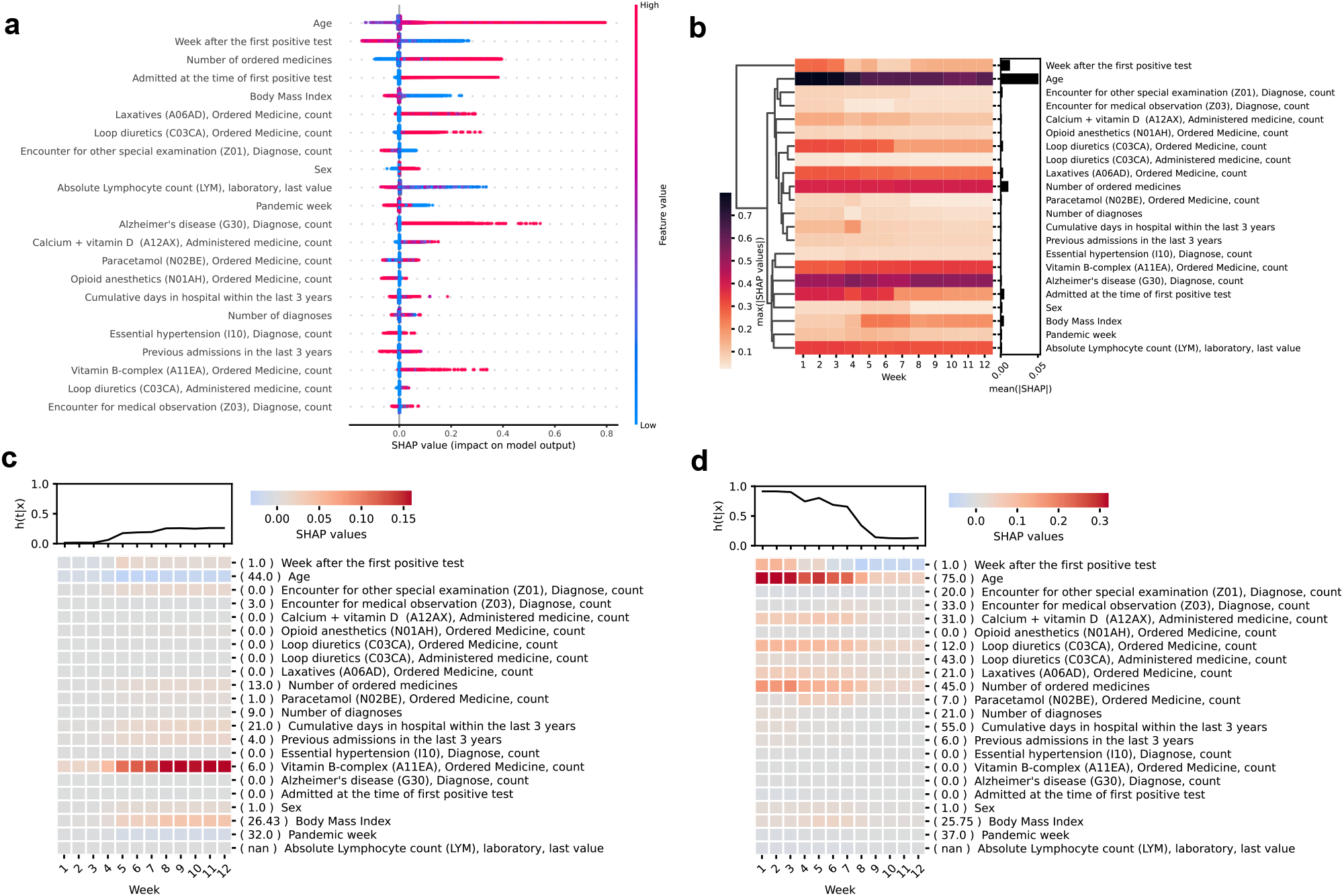
Global and local explanations of feature contributions to the risk of death in SARS-CoV-2 positive patients. SHAP values for each patient-week in the test set were calculated to explain the contribution of features to the discrete probability of death. A beeswarm plot (**a**) was generated to agglomerate all individual SHAP values for each patient-week with features coloured according to their normalised feature values. To explore the temporal dynamics, heatmaps were generated to show the maximum feature importance represented as the max(|SHAP|) across all patients (**b**) for each predicted week. The total feature importance of each feature was calculated as the mean(|SHAP|) across all weeks and shown as a bar plot (**b**). To exemplify personalized explanations, SHAP values for two patients (**c-d**) were depicted as heatmaps with their corresponding predicted discrete probabilities of death on top. The original feature values for each patient were reported inside round brackets next to the feature names. In all heatmaps, features were ordered by hierarchical clustering of the original feature values using Pearson correlation as the distance metric and average linkage.

### Machine learning captures non-linear patterns of mortality risk factors

Partial dependency plots (PDP) showed that the model learned non-linear contributions to the risk of mortality. We found that age contributes to the risk of death over 60 years of age (Fig 5a). BMI seemed to explain a higher risk of mortality in patients with BMI lower than 30 (Fig 5b), and males presented a higher risk of mortality than females. (Fig 5c). A higher risk of death was also seen for patients with low lymphocyte count (Fig 5d). As expected, patients with more hospitalizations and longer cumulative admission days prior to FPT exhibited a higher risk of death (Fig 5e-f). Similarly, the previously mentioned contribution of being admitted in the hospital at the time of the FPT to the risk of death was observed (Fig 5g). We found that the number of ordered medicines was a better predictor of death than the number of diagnoses, showing non-linear patterns where patients with less than five ordered medications in the last year showed up to 10% less risk of death whereas some patients with more than 20 ordered medications had up to 40% higher risk of death (Fig. 5h).

**Figure 5.**
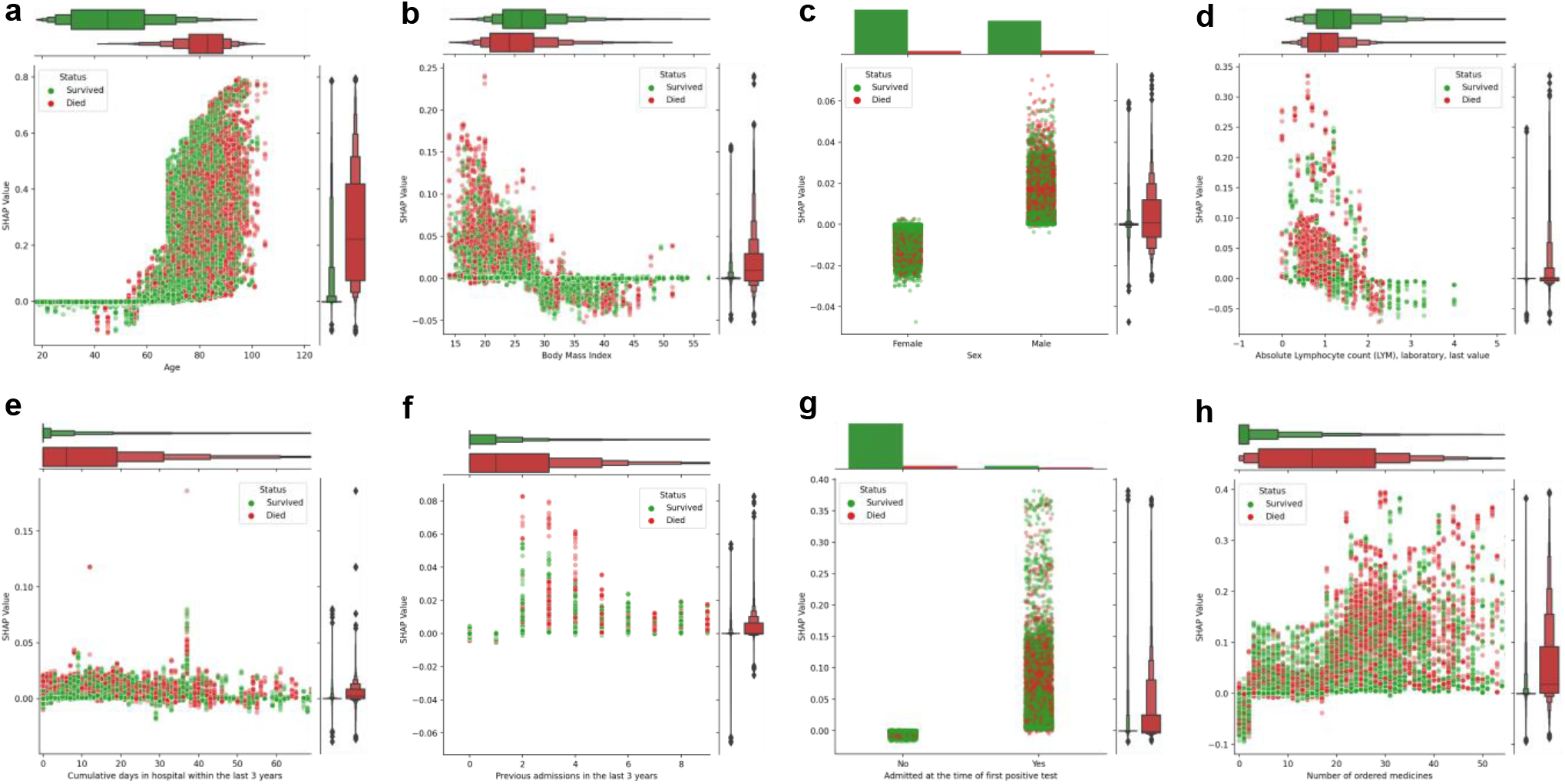
Individual feature explanations by survival status. Partial dependence plots (PDP) of SHAP values versus age (**a**), body mass index (**b**), sex (**c**), Lymphocytes levels (**d**), cumulative days in hospital (**e**) and the number of admissions (**f**) in the last 3 years, admission status at the time of first positive test (**g**) and the number of ordered medicines (**h**). Each dot shows a patient-week value coloured by survival status indicating those patients who survived (green) or died (red). Total SHAP values are represented as explained contributions in terms of probability (y-axis) given all the features values for a patient whereas features (x-axis) are represented by their corresponding value. The top and left panels of each PDP plot depict letter-value plots of the distribution of the x and y axes by survival status. Top panels were substituted by bar plots for categorical variables. Additional PDPs for the remaining features can be found in Supplementary Fig 2-4.

### Interactions between mortality risk factors reveal clusters of features

To unravel interactions between risk factors, we explored the interdependence of the selected features by their SHAP interactions values (Fig. 6). The interaction map for patients who died within 4 weeks from FPT revealed that the week of prediction feature and age interacted with several other features including previous hospital admissions and prescriptions of several drugs for at least 80% of patients (Fig. 6a). Thus, the information provided by these specific variables combined seems of particular importance for predicting early death (< 4 weeks). For patients who died after 8 weeks post-FPT, different interaction clusters emerged in which age, number of ordered medicines, BMI, and vitamin supplements like B-vitamins and calcium with vitamin D interacted in more than 70% of the patients. Also, lymphocyte count and admission at the time of FPT interacted with the number of medications in at least 60% of the patients (Fig. 6b).

**Figure 6.**
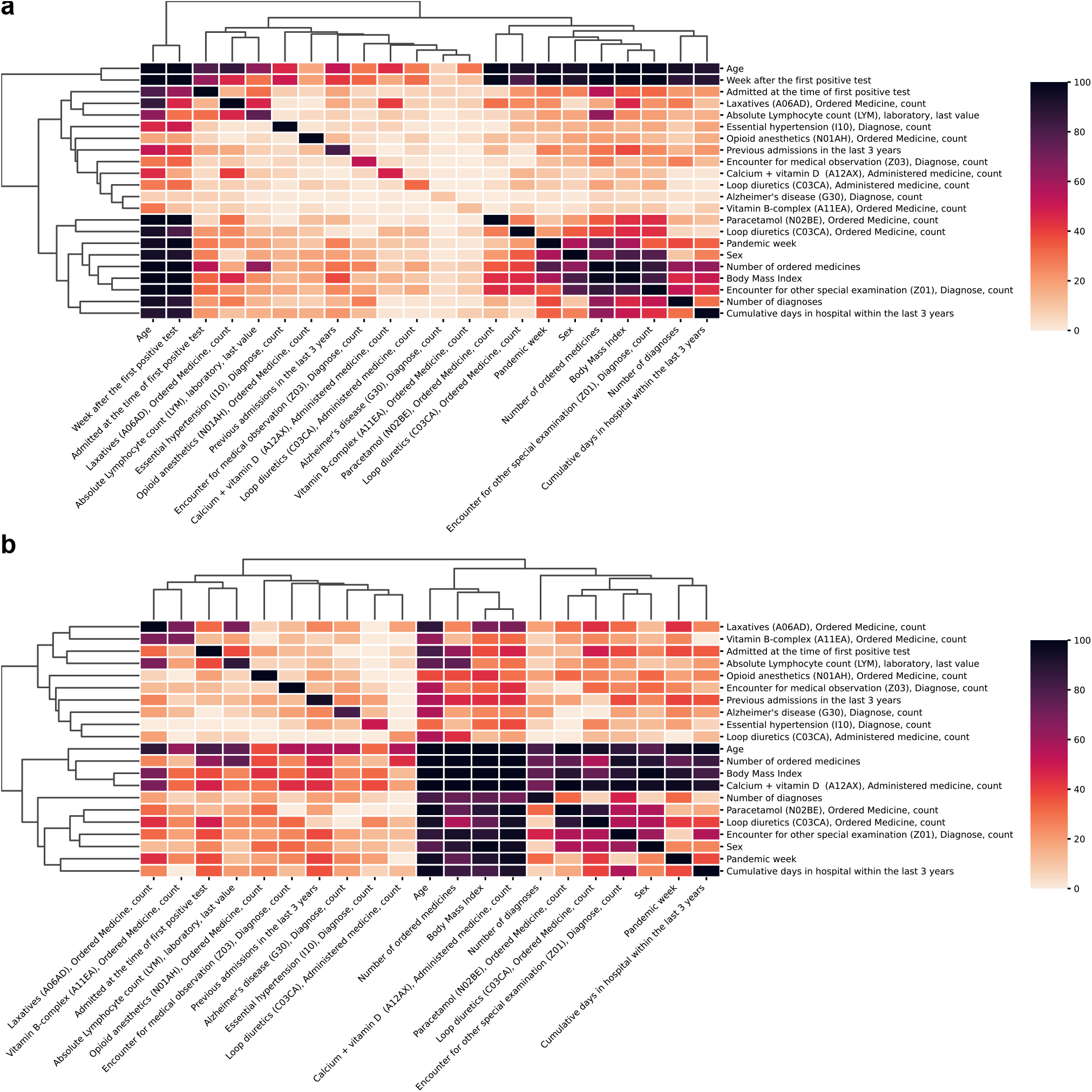
Summary of relevant feature interactions in explaining early and late mortality in SARS-CoV-2 positive patients. For each patient that died within 12 weeks, the SHAP interaction values between all 22 features were calculated. Only interaction values with an absolute value greater than 0.01 were considered relevant and counted. Counts were averaged across all patients to show the percentage rate a given pair of features was relevant. The diagonal represents the percentage of patients for which each feature had a SHAP value higher than 0.01. **a**, shows relevant feature interactions for patients who died within 4 weeks and for those who died between 8-12 weeks (**b**) – thus visualizing the difference in feature interactions for early and late mortality in SARS-CoV-2 positive patients. In both heatmaps, features were ordered by hierarchical clustering using Euclidean distance as the metric for average linkage.

## DISCUSSION

We here developed an explainable Machine Learning model for predicting the risk of death within the first 12 weeks from a positive SARS-CoV-2 PCR test. By implementing a discrete-time modelling approach we computed personalized survival probabilities, explained individual risk factors and achieved high discriminative performance in terms of C-index (0.946 CI 95%: 0.941-0.950) and PR-AUC (0.686 CI 95%: 0.651-0.720). Compared to traditional approaches we could model non-linear effects, learn interactions and explain temporal dynamics of risk factors without compromising discriminative performance.

During the COVID-19 pandemic, attempts have been made to provide prognostic models by implementing diverse modelling approaches. This has resulted in publications using statistical and Machine Learning (ML) approaches to predict the diagnosis or prognosis of COVID-19 related outcomes. Meta-analyses have indicated that the majority of published models suffer from a risk of bias due to overfitting, small sample sizes, poor cohort definition or not considering censored patients^18,29^. To overcome some of these previous limitations, we used electronic health records (EHR) from eastern Denmark, identifying 33,938 patients who had at least one positive SARS-CoV-2 RT-PCR test. To enable ML algorithms, clinical data need to be encoded into features that can be computed. Multiple approaches have been suggested for encoding EHR into computationally meaningful representations^30,31^. We opted for a simple feature engineering approach by considering the latest values or counts in clinically relevant time windows prior to FPT depending on the type of variable. Additionally, instead of characterizing patients’ relevant history using a limited set of pre-selected variables, the set of 22 features in the final model were derived using a data-driven approach from an initial set of 2,723 features that encoded available demographics, laboratory test results, hospitalizations, vital parameters, diagnoses and medicines. This approach enabled us to reduce model complexity to a smaller feature set while avoiding potential bias introduced by pre-selecting variables. While EHR are more representative of patient populations in terms of real-world data (RWD)^32^, some challenges arise when processing EHR for clinical research. Data collected from routine care may present inconsistencies^33^ that cannot be appropriately curated for in such big data sets, especially for information regarding clinical interventions or hospitalization status. We thus selected SARS-CoV-2 positive status and mortality for patient selection and outcome, respectively, based on robustness to bias from clinical management. Characteristics of these variables have been previously defined in a Danish nationwide cohort^28^ from 20^th^ of February 2020 until 19^th^ of May 2020 in alignment with our subset of patients in eastern Denmark.

More importantly, handling time in ML is not only relevant for encoding features but also for the modelling framework to use. When handling longitudinal data, time is usually fixed for a specific period and ML algorithms for binary classification are applied. To do so, patients for which the event of interest was not observed before they were lost to follow-up (censored) are excluded, resulting in underestimation of predicted risks^23,24^. This has been the predominant modelling approach in COVID-19^18,34^ related outcomes. Cox models^35^ are the most common statistical model for time-to-event considering censoring, but multiple ML algorithms allowing for censoring have been proposed^25^. Models such as regularized Cox models or Random Survival Forests have been successfully implemented for EHR^36^ and COVID-19^37^ data. These models are based on underlying assumptions such as proportional hazards in the case of Cox based models^35^ and handle time as continuous. An alternative is to consider time as discrete^26,35^ which has demonstrated performance as good or better than continuous-time models^38,39^ with the advantage of accounting for censoring while enabling the implementation of existing ML algorithms such as gradient boosting decision trees^40^. In this way, we overcame the limitations of Cox based models, by training ML models that learned complex interactions and non-linear effects from the data. Moreover, because no proportionality of hazards was assumed, our model could predict personalized survival probabilities^27^ for each patient given their specific context, further facilitating a precision medicine approach^41^.

To understand model predictions, ML explainability, or explainable artificial intelligence (xAI), is particularly powerful to enable scientific insights by leveraging the ability of ML models to learn complexity transcending traditional assumptions^21^. In some cases, seemingly paradoxical effects have been unraveled when modelling clinical data^42^. Multiple approaches have been proposed to open “black-box” models and allow explainability by, for example, removing features and measuring their impact on the model^43^. These methods have been successfully applied in clinical research for various diseases^20,44^, but in the case of COVID-19^45^ most of these are limited to scenarios of binary classification that ignored censoring. As an alternative approach, we provide explanations of the model predictions based on SHAP values^46^ that not only decompose the predicted survival probability for each patient in terms of the features’ contributions but also reflect temporal dynamics of such contributions in the context of time-to-event modelling. Local explanations as provided in our study are critical for precision medicine by indicating patient-specific risk factors, but also raise epistemological challenges on how to extrapolate from local to global explanations^47,48^. We employed traditional summary statistics to shed some light on common risk factors, but such reduction of complexity may imply a reduction of granularity of factors that are not relevant at the population level but critical for specific patients. Importantly, the features selected as good predictors do not necessarily imply causality^21,52^ and different sets of features have been demonstrated to be equally predictive in terms of performance in some cases^49^.

In line with previous studies, we here identified high age^15^ and sex (male)^50^ as important risk factors in COVID-19. As the importance of age increased significantly for age over 60 years, while capturing high age as a risk factor in itself, our model may further reflect other age-related factors such as an increased prevalence of comorbidities, which is supported by the interaction plots. BMI and obesity have previously been reported as risk factors for severe COVID-19^13^ and severe obesity as a risk factor for COVID-related mortality, especially for younger patients^51^, who are likely candidates for ICU care and treatment with mechanical ventilation, resulting in improved survival. In contrast, we identified an increased risk of death for patients with BMI below 30. This could reflect several other risk factors associated with low BMI, such as elderly, frail, patients with comorbidity. This is supported by the interaction plots demonstrating an interaction between BMI and the number of ordered medicines in early deaths, and interactions with the number of diagnoses, cumulative days in hospital prior to FPT, and several specific medications for late deaths. Lymphocytopenia was also identified as a predictor of high mortality in line with previous findings^52^. This may be a proxy for immune dysfunction, due to prior or ongoing therapy, malignancy or comorbidity, as well as a severe ongoing COVID-19 disease itself.

As expected, an increased risk of death was observed in patients with an increased number of medications and diagnosis codes, likely representing comorbidities, in line with previous studies^53^. We found that the number of ordered medicines was a better predictor of death than the number of diagnoses, emphasizing the need to capture disease burden based on actual medication in addition to coded diagnoses. This highlights the need to further explore feature encoding of clinical variables^30^, to more accurately represent clinical concepts such as comorbidities. We also observed that hospital encounters for medical examination with known or unknown causes correlated with a lower risk of death. This may indicate in-patient management of COVID-19 early in the pandemic or reflect increased monitoring of patients with anticipated increased risk of COVID-19, thereby enabling earlier interventions. Similarly, including the pandemic week in which a patient had their FPT as a feature revealed that patients early rather than later in the pandemic, had a higher risk of dying. As our data covered both the first and second pandemic wave in Denmark, this finding likely reflects that our model captured improvements in the clinical management of patients throughout the pandemic^54^.

The implemented discrete-time modelling approach required encoding the week from FPT as a feature, revealing explanations of temporal dynamics through SHAP values. When interpreting this feature, a higher risk of death in the first four weeks was observed, probably capturing the risk due to active infection during that period^55^. Critically, our model could differentiate between risk factors for early vs late mortality. Here, hospital admission at the time of FPT, pandemic week of prediction, age and ordering of loop diuretics were important factors for early death. Meanwhile, factors explaining the risk of late death (>8 weeks) included lower BMI as a potential proxy for frail patients, diagnosis of Alzheimer’s disease, and ordered B-vitamin (a probable indicator of patient malnutrition or alcohol abuse). These factors likely represent patient groups who may not respond well to treatment and are likely not candidates for ICU or mechanical ventilation, thus exhibiting disease progression leading to late mortality. This is supported by the interactions observed between age, number of ordered medicines, (low) BMI, and various vitamin supplements, which are factors likely reflecting patient frailty. Interestingly, age and number of medicines as a proxy for comorbidity burden before SARS-CoV-2 infection remained prominent risk factors throughout the disease course. This suggests that predicting late deaths requires a different set of risk factors and consideration of their interactions than predicting early death. Thus, uncovering the interdependency of features important for early vs late death also indicated time dependency of risk factors.

## CONCLUSION

We developed a data-driven machine learning model to identify SARS-CoV-2 positive patients with a high risk of death within 12-week from the first positive test. The discrete-time modelling approach implemented not only allowed us to train survival models with high performance but also enabled model explainability through SHAP values. By learning temporal dynamics and interactions between clinical features, the model was able to identify personalized risk factors and high-risk patients for early interventions while improving the understanding of the disease. At the same time, we demonstrate that leveraging electronic health records with explainable ML models provide a framework for the implementation of precision medicine in routine care which can be adapted to other diseases.

## METHODS

### Data sources

The study is approved by the Danish Regional Ethical Committee (H-20026502) and Data Protection Agency (P-2020-426). Data were obtained retrospectively from raw electronic health records (EHR) from the Capital Region and Region Zealand (eastern Denmark), covering a population of 2,761,556 people. Data from the electronic patient journal (EPJ) by EPIC systems, is logged and stored in the Chronicles database containing live and historic data. Daily extracts are transferred into the Clarity and Caboodle databases. The final dataset was extracted from the Caboodle database containing data up to the 2^nd^ of March 2021. Real-Time Polymerase Chain Reaction (RT-PCR) SARS-CoV-2 test results were used to identify 963,265 individuals over 18 years old with a test taken between the 17th of March 2020 and 2^nd^ of March 2021 in eastern Denmark.

### Feature engineering

Features were generated according to different data types and retrospective time windows including observations until the day of the first positive SARS-CoV-2 test (FPT). Basic characteristics such as age, sex, and body mass index (BMI) were encoded as the latest value observed up to the day of FPT. Measurements represented as continuous values such as laboratory test results (e.g. lymphocyte levels) and vital parameters (e.g. systolic blood pressure) were encoded as the latest value observed in the last month before the FPT. For variables measured as categorical values represented by domain specific codes, features were generated by counting the total number of occurrences of each and all codes in defined time windows. For diagnoses represented by International Statistical Classification of Diseases and Related Health Problems version 10 (ICD-10) codes, the selected time window was three years, while for medications represented by Anatomical Therapeutic Chemical (ATC) codes, the time window was one year. Previous hospitalisations, defined as hospital stays longer than 24h, were encoded as cumulative days in hospital within the last three years as well as the total count of hospital admissions in this time period. Features that may help guide the algorithm by providing a context of external events were also included. Among these features, the number of weeks since the start of the pandemic until the FPT was taken and a binary feature indicating if the patient was hospitalized when the FPT occurred. Missingness was assumed to be informative and not at random. For diagnoses and medications, the lack of a code was assumed to be not assigned and encoded as a zero in the features. For continuous variables such as laboratory values and vitals, missingness was accounted for by the tree-based ML algorithm chosen without the need for imputation.

### Machine Learning approach to survival modelling

To perform time-to-event modelling we considered a discrete-time modelling approach^26^ to predict 12-week mortality since a first SARS-CoV-2 positive test. Described by Cox as an approximation to his proposed proportional hazards assumption for continuous-time modelling^35^, discretizing time in intervals allowed us, to perform binary classification at each time interval. By doing this, we trained models that accounted for right-censored observations, hence reducing the risk of selection bias^23^, and estimated conditional probabilities of death given the features that could be computed and explained efficiently without stringent assumptions. Data was generated from EHR on 2^nd^ of March 2021, hence right-censoring was observed for patients that had a positive test from 8^th^ of December 2020 (12-weeks before data generation) and did not die. The survival status of these patients could not be ascertained in such a period hence they were only considered for the follow-up period available. Deaths that occurred the same day of the first positive test (FPT) were excluded. During the training phase, the original dataset was augmented longitudinally by repeating each patient’s feature set containing values up to the FPT into patient-weeks. The feature vector for a patient was repeated according to the number of weeks since the FPT up to the week of death or censoring for a maximum of 12 weeks since FPT. The main difference between each row is that time was encoded as an ordinal feature indicating the week of prediction with values ranging from 1 to 12. The target values for each patient-week were set to 0 up to the week of death or censoring which were indicated as a 1 or a 0 respectively. When using the trained models for prediction, the feature set with values up to the FPT for each patient was augmented longitudinally 12 times. Time was encoded as an ordinal feature with values 1 to 12 so 12 probabilities of death, one probability per week per patient, would be predicted. The predicted probabilities of death constitute the hazard function *h*(*t*|*x*) which can be also expressed as a survival function *S*(*t*|*x*) and a cumulative density function *F*(*t*|*x*) as defined below:

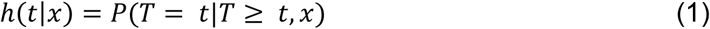

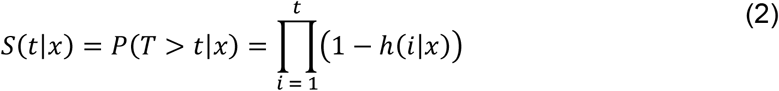

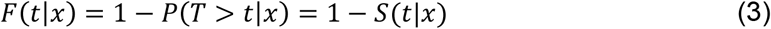

### Local and global explainability

SHAP values^22^ were calculated to quantify the local contribution of each feature to the risk of death of each individual at each predicted week. Based on Shapley values originally described in the context of game theory, SHAP values were computed exactly and efficiently for our tree-based models using TreeSHAP^56^. The SHAP values computed in log-odds space for all models trained in the ensemble were averaged and transformed into probabilities by linear scaling. These probabilities represent the local contribution of each feature to the hazard h(*t*|*x*) for each predicted week. Similarly, SHAP interaction values were calculated to assess the contribution of each pair of features as previously described^46^. Local SHAP interaction values can be understood as the difference between SHAP values for each pair of features when one of the features is not present.

While local interpretations are useful to understand patient-specific risk factors, global explanations can reveal general risk factors by summarizing local explanations. To do so, SHAP values were used to estimate feature importance. We computed each feature’s importance in terms of absolute and mean SHAP values for each feature. Feature selection was performed by removing features with a mean(|SHAP|) < 0.01. Both local and global interpretations were provided to clinicians for generating clinical explanations of the risk factors.

### Model development and assessment

We trained gradient boosting decision tree models (LightGBM^40^) using cross-entropy as the objective function for optimization. To do this, the full dataset was split into training (60%), validation (20%), and test (20%) sets each one with the same distribution of deaths. Cross-validation (CV) was performed in two steps. First, the training set was divided into 5 subsets and the subsample rate (0.7), learning rate (0.05), number of iterations (50) and positive class weight (100) were adjusted using 5-fold cross-validation while the rest of the parameters were set to default (Supplementary table 3). Once suitable parameters were found, feature selection was performed based on the validation set. Second, the training set and validation set were combined and split into 5 folds to re-train and generate a final ensemble of 5 models trained on 80% of the data. The performance reported was assessed by averaging the predictions of the ensemble on the test set (20%), which was not used for model development.

Based on the predicted cumulative probabilities of death, time-to-event performance was measured by the concordance index (C-index) based on the inverse probability of censoring weights^57^ across all weeks. Performance was further assessed at each week by excluding right-censored cases when calculating binary metrics and measured in terms of precision-recall area under the curve (PR-AUC), Mathew Correlation Coefficient (MCC)^58^, sensitivity and specificity. A threshold of 0.5 was used to turn predicted probabilities into binary classes. Confidence intervals (95% CI) for the performance metrics were calculated by bootstrapping with resampling for 1000 iterations.

### Software

Data wrangling was performed using R^59^ and the tidyverse library^60^. Feature engineering was performed in Python using the pandas^61^ and numpy^62^ libraries. Gradient boosting decision trees were trained and implemented using LightGBM^40^ assessing model performance using the implementations in scikit-Learn^63^ and scikit-survival^64^. Summary statistics were generated using tableone^65^.

## Data Availability

Data can be requested through the corresponding author, however, due to data protection regulations, data cannot be made publicly available, but the authors will assist external researchers in accessing the data on a collaborative basis upon request.

## DATA AND CODE AVAILABILITY

Data can be requested through the corresponding author, however, due to data protection regulations, data cannot be made publicly available, but the authors will assist external researchers in accessing the data on a collaborative basis upon request. The trained models and code to run predictions are publicly available on Github under a GNU Affero General Public License v3.0 (https://github.com/PERSIMUNE/COVIMUN_DT)

## ACKNOWLEDGEMENTS

The study was supported by a COVID-19 grant from the Ministry of Higher Education and Science (0238-00006B) and the Danish National Research Foundation (DNRF126). The Capital Region of Denmark, Center for Economy, provided data extracts from the EHR system.

## CONTRIBUTIONS

A.G.Z, R.A, S.R.O and C.U.N conceived the project and supervised it. A.G.Z, R.A and K.S.M. performed data cleaning. A.G.Z and R.A developed the model and visualizations. A.G.Z, R.A, S.R.O, C.U.N. designed the study. A.G.Z, R.A, R.S, K.S.M., R.Z.M, S.R.O, C.U.N. interpreted the data and results. A.G.Z, R.A, R.S, R.Z.M, C.U.N. wrote the paper. All authors commented on and approved the final manuscript.

