## Supplementary figure and tables for "Personalized survival probabilities for SARS-CoV-2 positive patients by explainable machine learning"

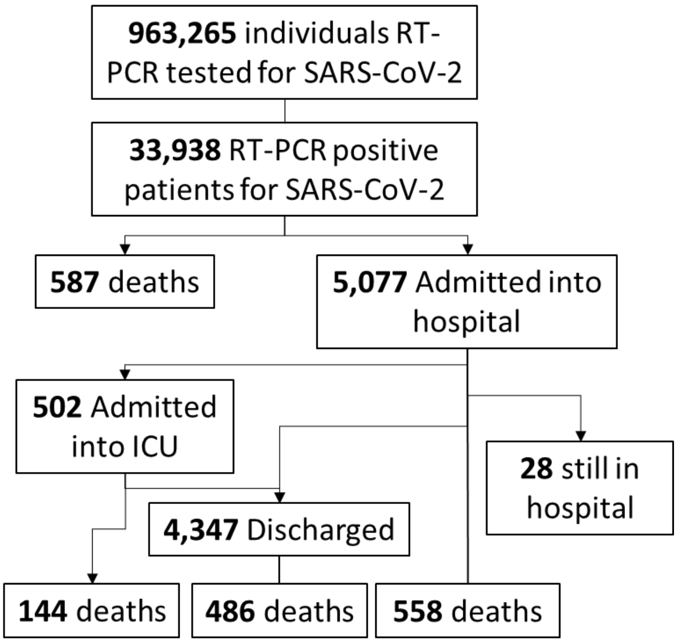


### Supplementary figure 1. Consort diagram of the cohort.

963,265 individuals were identified using Real-Time Polymerase Chain Reaction (RT-PCR) SARS-CoV-2 test results taken between the 17th of March 2020 and 2nd of March 2021 in eastern Denmark. All reported numbers correspond to positive tests, admissions, discharges and deaths that occurred in the mentioned period independently if such events occurred after 12 weeks from a first positive test.

| Group | TP | FN | FP | TN | Precision | Sensitivity | Specíficity | ROC-AUC | Censored | Deaths |
|---|---|---|---|---|---|---|---|---|---|---|
| **All patients** | 552 | 4 | 851 | 5408 | 0.3934 | 0.9928 | 0.864 | 0.9703 | 5359 | 556 |
| **Patients tested outside the hospital** | 371 | 4 | 592 | 5292 | 0.3853 | 0.9893 | 0.8994 | 0.9774 | 5022 | 375 |
| **Patients admitted to the hospital at the time of test** | 181 | 0 | 259 | 116 | 0.4114 | 1 | 0.3093 | 0.8576 | 337 | 181 |

### Supplementary Table 1. Binary performance metrics by admission status.

Binary metrics were assessed 12 weeks from the first positive test. True positives (TP), false negatives (FN), false positives (FP), true negatives (TN), area under the receiver operating characteristic curve (ROC-AUC).

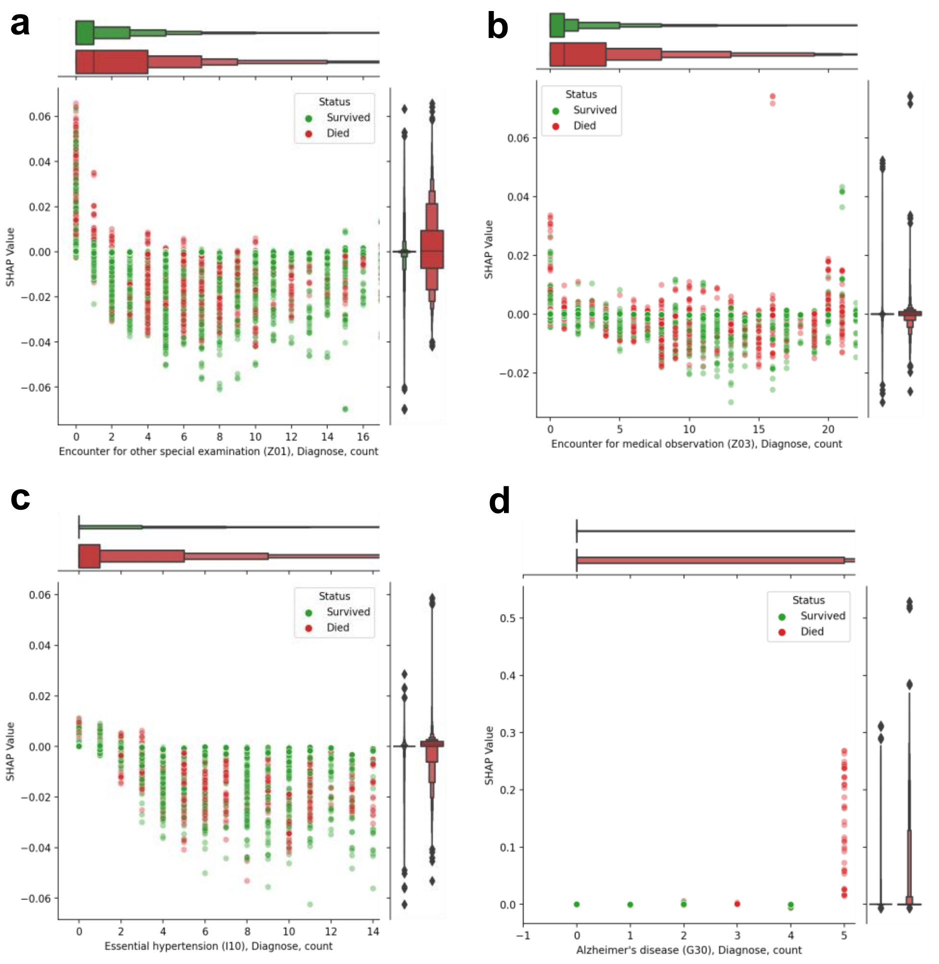


**Supplementary Figure 2. Partial dependence plots of diagnoses by survival status.**

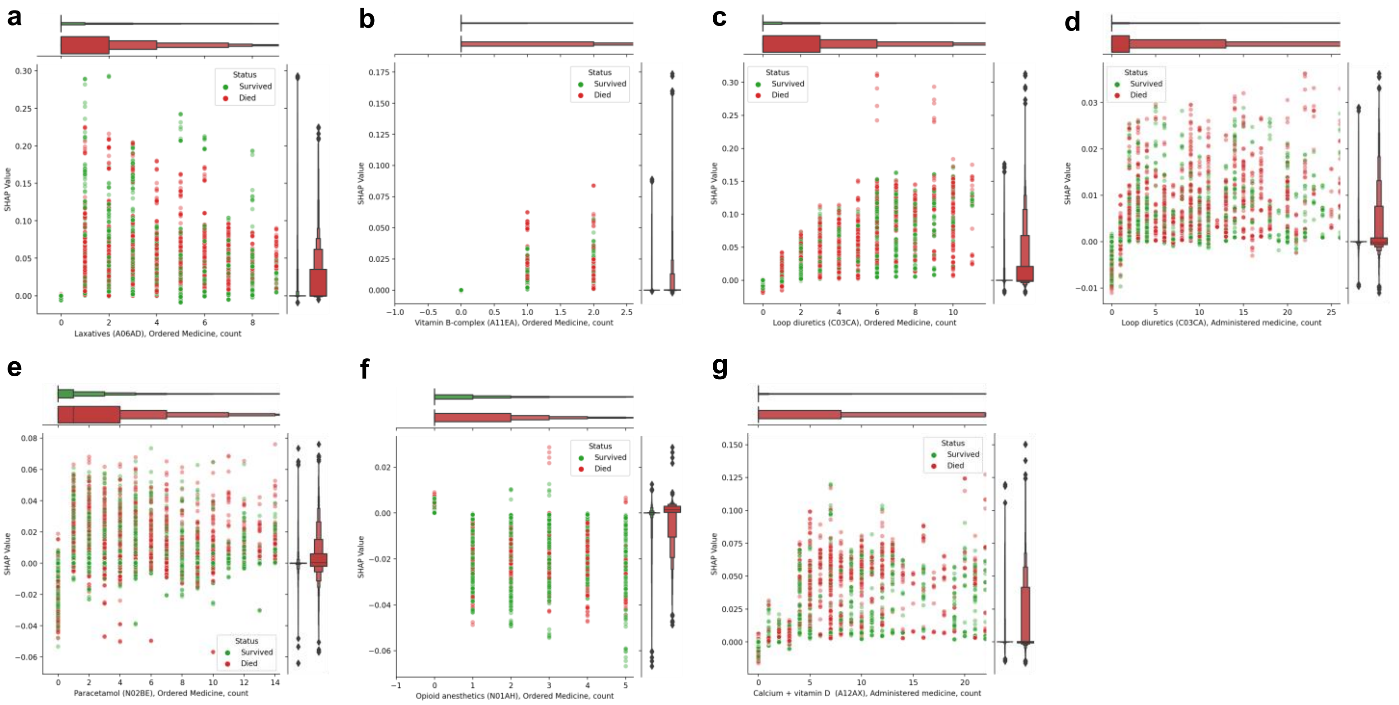


**Supplementary Figure 3. Partial dependence plots of medications by survival status.**

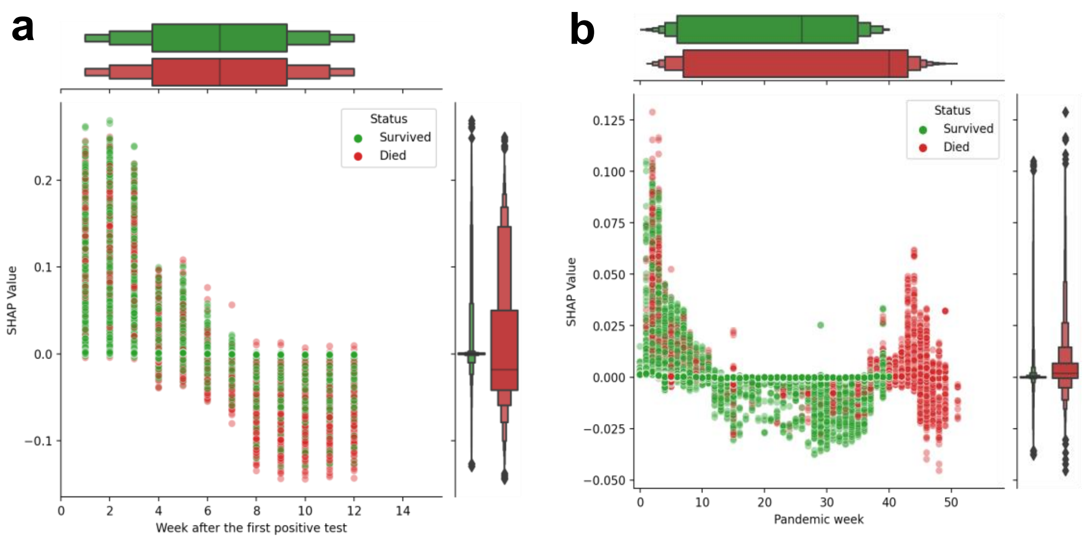


**Supplementary Figure 4. Partial dependence plots of temporal features by survival status.**

**Diagnoses represented as ICD -10 codes**

A01, A02, A04, A05, A06, A07, A08, A09, A15, A16, A17, A18, A23, A24, A26, A28, A35, A37, A38, A40, A41, A44, A46, A48, A49, A51, A52, A53, A54, A56, A60, A63, A64, A68, A69, A70, A74, A77, A79, A80, A81, A86, A87, A88, A89, B00, B01, B02, B07, B08, B09, B15, B16, B17, B18, B20, B22, B23, B25, B26, B27, B30, B33, B34, B35, B36, B37, B44, B48, B49, B50, B51, B54, B55, B58, B65, B66, B67, B71, B76, B80, B83, B86, B90, B91, B94, B95, B96, B97, B98, B99, C01, C02, C04, C05, C06, C07, C09, C10, C11, C13, C15, C16, C17, C18, C20, C21, C22, C23, C24, C25, C26, C30, C32, C34, C37, C38, C40, C43, C44, C45, C46, C48, C49, C50, C51, C52, C53, C54, C56, C57, C60, C61, C62, C64, C65, C66, C67, C69, C71, C73, C74, C76, C77, C78, C79, C80, C81, C82, C83, C84, C85, C86, C88, C90, C91, C92, C93, C94, C95, C96, C99, D03, D04, D05, D06, D07, D09, D10, D11, D12, D13, D14, D15, D16, D17, D18, D19, D20, D21, D22, D23, D24, D25, D27, D28, D29, D30, D31, D32, D33, D34, D35, D36, D37, D38, D39, D40, D41, D43, D44, D45, D46, D47, D48, D50, D51, D52, D55, D56, D58, D59, D60, D61, D62, D63, D64, D65, D66, D67, D68, D69, D70, D71, D72, D73, D74, D75, D76, D80, D82, D83, D84, D86, D89, E03, E04, E05, E06, E07, E10, E11, E13, E14, E15, E16, E20, E21, E22, E23, E24, E25, E26, E27, E28, E29, E30, E31, E34, E41, E46, E47, E50, E51, E53, E55, E56, E58, E60, E61, E63, E64, E65, E66, E67, E68, E70, E72, E73, E74, E75, E78, E79, E80, E83, E84, E85, E86, E87, E88, E89, F00, F01, F02, F03, F04, F05, F06, F07, F09, F10, F11, F12, F13, F14, F15, F16, F17, F19, F20, F21, F22, F23, F25, F28, F29, F30, F31, F32, F33, F34, F38, F39, F40, F41, F42, F43, F44, F45, F48, F50, F51, F52, F53, F55, F59, F60, F61, F62, F63, F64, F65, F68, F70, F71, F72, F73, F78, F79, F80, F81, F82, F84, F88, F89, F90, F91, F92, F93, F94, F95, F98, F99, G00, G02, G03, G04, G05, G06, G10, G11, G12, G14, G20, G21, G22, G23, G24, G25, G30, G31, G35, G36, G37, G40, G41, G43, G44, G45, G46, G47, G50, G51, G52, G53, G54, G55, G56, G57, G58, G59, G60, G61, G62, G63, G64, G70, G71, G72, G73, G80, G81, G82, G83, G90, G91, G92, G93, G94, G95, G96, G97, G98, G99, H00, H01, H02, H03, H04, H05, H06, H10, H11, H13, H15, H16, H17, H18, H19, H20, H21, H22, H25, H26, H27, H28, H30, H31, H33, H34, H35, H36, H40, H42, H43, H44, H45, H46, H47, H48, H49, H50, H51, H52, H53, H54, H55, H57, H58, H60, H61, H62, H65, H66, H68, H69, H70, H71, H72, H73, H74, H80, H81, H82, H83, H90, H91, H92, H93, H94, H95, I05, I06, I07, I10, I11, I12, I13, I15, I20, I21, I23, I24, I25, I26, I27, I30, I31, I32, I33, I34, I35, I36, I37, I38, I39, I40, I42, I44, I45, I46, I47, I48, I49, I50, I51, I52, I60, I61, I62, I63, I64, I65, I66, I67, I68, I69, I70, I71, I72, I73, I74, I77, I78, I79, I80, I81, I82, I83, I85, I86, I87, I88, I89, I95, I97, I99, J00, J01, J02, J03, J04, J05, J06, J09, J10, J11, J12, J13, J14, J15, J16, J17, J18, J20, J21, J22, J30, J31, J32, J33, J34, J35, J36, J37, J38, J39, J40, J41, J42, J43, J44, J45, J46, J47, J61, J62, J64, J67, J68, J69, J70, J80, J81, J82, J84, J85, J86, J90, J91, J92, J93, J94, J95, J96, J98, K00, K01, K02, K03, K04, K05, K06, K07, K08, K09, K10, K11, K12, K13, K14, K20, K21, K22, K25, K26, K27, K28, K29, K30, K31, K35, K36, K37, K38, K40, K41, K42, K43, K44, K45, K46, K50, K51, K52, K55, K56, K57, K58, K59, K60, K61, K62, K63, K64, K65, K66, K70, K71, K72, K73, K74, K75, K76, K80, K81, K82, K83, K85, K86, K87, K90, K91, K92, L00, L01, L02, L03, L04, L05, L08, L10, L11, L12, L13, L20, L21, L22, L23, L24, L25, L26, L27, L28, L29, L30, L40, L41, L42, L43, L44, L50, L51, L52, L53, L55, L56, L57, L58, L59, L60, L63, L64, L65, L66, L67, L68, L70, L71, L72, L73, L74, L80, L81, L82, L84, L85, L88, L89, L90, L91, L92, L93, L94, L95, L97, L98, L99, M00, M02, M05, M06, M07, M08, M10, M11, M12, M13, M14, M15, M16, M17, M18, M19, M20, M21, M22, M23, M24, M25, M30, M31,

| |
|---|
| M32, M33, M34, M35, M40, M41, M42, M43, M45, M46, M47, M48, M49, M50, M51, M53, M54, M60, M61, M62, M65, M66, M67, M68, M70, M71, M72, M75, M76, M77, M79, M80, M81, M82, M83, M84, M85, M86, M87, M88, M89, M90, M91, M92, M93, M94, M95, M96, M99, N00, N02, N03, N04, N05, N06, N08, N10, N11, N12, N13, N15, N16, N17, N18, N19, N20, N21, N25, N26, N27, N28, N30, N31, N32, N34, N35, N36, N39, N40, N41, N42, N43, N44, N45, N46, N47, N48, N49, N50, N51, N60, N61, N62, N63, N64, N70, N71, N72, N73, N74, N75, N76, N80, N81, N82, N83, N84, N85, N86, N87, N88, N89, N90, N91, N92, N93, N94, N95, N96, N97, N98, N99, O00, O01, O02, O03, O04, O05, O07, O08, O10, O12, O13, O14, O16, O20, O21, O22, O23, O24, O26, O28, O30, O31, O32, O34, O35, O36, O40, O41, O42, O43, O44, O45, O46, O47, O48, O49, O60, O61, O62, O63, O64, O65, O66, O67, O68, O69, O70, O71, O72, O73, O74, O75, O80, O81, O82, O83, O84, O85, O86, O87, O88, O89, O90, O91, O92, O98, O99, P01, P02, P05, P20, P29, P74, P92, Q00, Q03, Q04, Q05, Q06, Q07, Q10, Q11, Q12, Q14, Q15, Q17, Q18, Q20, Q21, Q22, Q23, Q24, Q25, Q26, Q27, Q28, Q30, Q32, Q35, Q36, Q37, Q39, Q40, Q43, Q51, Q53, Q54, Q61, Q62, Q63, Q64, Q65, Q66, Q67, Q68, Q72, Q74, Q75, Q77, Q78, Q79, Q81, Q82, Q83, Q84, Q85, Q86, Q87, Q89, Q90, Q91, Q92, Q93, Q95, Q96, Q97, Q98, Q99, R00, R01, R02, R03, R04, R05, R06, R07, R09, R10, R11, R12, R13, R14, R15, R16, R17, R18, R19, R20, R21, R22, R23, R25, R26, R27, R29, R30, R31, R32, R33, R34, R35, R39, R40, R41, R42, R43, R44, R45, R46, R47, R48, R49, R50, R51, R52, R53, R55, R56, R57, R58, R59, R60, R61, R62, R63, R64, R67, R68, R69, R70, R71, R73, R74, R76, R77, R78, R79, R80, R81, R82, R84, R87, R89, R90, R91, R92, R93, R94, S00, S01, S02, S03, S04, S05, S06, S07, S09, S10, S11, S12, S13, S14, S15, S19, S20, S21, S22, S23, S24, S25, S27, S29, S30, S31, S32, S33, S34, S36, S37, S38, S39, S40, S41, S42, S43, S44, S45, S46, S49, S50, S51, S52, S53, S54, S56, S57, S59, S60, S61, S62, S63, S64, S65, S66, S67, S68, S69, S70, S71, S72, S73, S74, S76, S79, S80, S81, S82, S83, S84, S85, S86, S87, S89, S90, S91, S92, S93, S96, S97, S98, S99, T00, T01, T02, T04, T07, T08, T09, T10, T11, T12, T13, T14, T15, T16, T17, T18, T19, T20, T21, T22, T23, T24, T25, T26, T27, T28, T29, T30, T31, T32, T33, T35, T38, T39, T40, T41, T42, T43, T45, T46, T47, T50, T51, T52, T53, T54, T55, T58, T59, T62, T63, T65, T66, T67, T68, T69, T70, T71, T73, T74, T75, T78, T79, T80, T81, T82, T83, T84, T85, T86, T87, T88, T90, T91, T92, T93, T95, T98, V03, V10, V11, VRA, VRB, VRK, X60, X61, X62, X63, X64, X69, X70, X71, X78, X81, X82, X83, X84, X91, X95, X99, Y04, Z00, Z01, Z02, Z03, Z04, Z06, Z07, Z08, Z09, Z10, Z11, Z12, Z13, Z20, Z21, Z22, Z23, Z24, Z25, Z26, Z27, Z29, Z30, Z31, Z32, Z34, Z35, Z36, Z37, Z38, Z39, Z40, Z41, Z42, Z47, Z48, Z50, Z51, Z52, Z54, Z55, Z56, Z57, Z58, Z59, Z60, Z61, Z62, Z63, Z64, Z65, Z70, Z71, Z72, Z73, Z74, Z75, Z76, Z80, Z81, Z82, Z83, Z84, Z85, Z86, Z87, Z88, Z89, Z90, Z91, Z92, Z93, Z94, Z95, Z96, Z97, Z98, Z99 |
| **Medications represented as ATC codes**<br>A01*, A01AA, A01AB, A01AC, A01AD, A02AA, A02AD, A02AH, A02BA, A02BB, A02BC, A02BX, A02X, A03AA, A03AB, A03AX, A03BA, A03BB, A03FA, A04AA, A04AD, A05AA, A05BA, A06AA, A06AB, A06AC, A06AD, A06AG, A06AH, A06AX, A07AA, A07BA, A07CA, A07DA, A07EA, A07EC, A07FA, A07XA, A08AA, A08AB, A09AA, A10AB, A10AC, A10AD, A10AE, A10BA, A10BB, A10BD, A10BH, A10BJ, A10BK, A10BX, A11AA, A11AB, A11CA, A11CC, A11DA, A11E, A11EA, A11EB, A11GA, A11HA, A11JC, A12*, A12A, A12AA, A12AX, A12BA, A12CA, A12CB, A12CC, A12CE, A12CX, A16AA, A16AX, B01*, B01AA, B01AB, B01AC, B01AD, B01AE, B01AF, B01AX, B02AA, B02BA, B02BB, B02BC, B02BD, B02BX, B03A, B03AA, B03AB, B03AC, B03AE, B03BA, B03BB, B03XA, B05*, B05AA, B05BA, B05BB, B05BC, B05CX, B05DA, B05DB, B05XA, B05XC, B06AC, C***, C01AA, C01BC, C01BD, C01CA, C01CE, C01CX, C01DA, C01DX, C01EB, C02AB, C02AC, C02CA, C02DB, C02DC, C02DD, C02KX, C03AA, C03AB, C03BA, C03CA, C03DA, C03EA, C03EB, C03XA, C05AA, C05AE, C05BA, C05BB, C05CA, C07AA, C07AB, C07AG, C07BB, C07CB, C08CA, C08DA, C08DB, C09AA, C09BA, C09CA, C09DA, C09DX, C09XA, C10AA, C10AB, C10AC, C10AD, C10AX, C10BA, D01AC, D01AE, D01BA, D02AB, D02AC, D02AE, D02AF, D02AX, D03AX, D04AB, D05AA, D05AX, D05BA, D05BB, D06AA, D06AX, D06BA, D06BB, D06BX, D07AA, D07AB, D07AC, D07AD, D07BC, D07CA, D07CC, D07XC, D08AB, D08AC, D08AJ, D08AX, D10AB, D10AD, D10AE, D10AF, D10AX, D10BA, D11A, D11AH, D11AX, G01*, G01AA, G01AF, G02AB, G02AD, G02BA, G02BB, G02CB, G02CX, G03AA, G03AB, G03AC, G03AD, G03BA, G03CA, G03CX, G03DA, G03DB, G03FA, G03FB, G03GA, G03GB, G03HA, G03HB, G03XB, G03XC, G04BD, G04BE, G04CA, G04CB, H01AA, H01AB, H01AC, H01AX, H01BA, H01BB, H01CA, H01CB, H01CC, H02AA, H02AB, H03AA, H03BA, H03BB, H03CA, H04AA, H05AA, H05BA, H05BX, J01AA, J01CA, J01CE, J01CF, J01CR, J01DB, J01DC, J01DD, J01DH, J01DI, J01EA, J01EB, J01EC, J01EE, J01FA, J01FF, J01GB, J01MA, J01XA, J01XB, J01XC, J01XD, J01XE, J01XX, J02AA, J02AC, J02AX, J04AB, J04AC, J04AK, J04AM, J04BA, J05AB, J05AD, J05AE, J05AF, J05AG, J05AH, J05AJ, J05AP, J05AR, J05AX, J06BA, J06BB, J07AE, J07AG, J07AH, J07AJ, J07AL, J07AM, J07AP, J07BA, J07BB, J07BC, J07BD, J07BF, J07BK, J07BM, L***, L01*, L01AA, L01AB, L01AC, L01AD, L01AX, L01BA, L01BB, L01BC, L01CA, L01CB, L01CD, L01CE, L01CX, L01DB, L01DC, L01EA, L01EB, L01EE, L01EJ, L01EL, L01EX, L01XA, L01XB, L01XC, L01XD, L01XE, L01XF, L01XG, L01XK, L01XX, L01XY, L02AB, L02AE, L02BA, L02BB, L02BG, L02BX, L03AA, L03AB, L03AX, L04*, L04AA, L04AB, L04AC, L04AD, L04AX, M01AA, M01AB, M01AC, M01AE, M01AG, M01AH, M01AX, M02AA, M03AB, M03AC, M03AX, M03BB, M03BX, M03CA, M04AA, M04AB, M04AC, M05BA, M05BB, M05BX, N01AF, N01AH, N01AX, N01BA, N01BB, N02AA, N02AB, N02AE, N02AG, N02AJ, N02AX, N02B, N02BA, N02BE, N02BG, N02CA, N02CC, N02CD, N02CX, N03AA, N03AB, N03AD, N03AE, N03AF, N03AG, N03AX, N04AA, N04AB, N04BA, N04BB, N04BC, N04BD, N04BX, N05AA, N05AB, N05AD, N05AE, N05AF, N05AG, N05AH, N05AL, N05AN, N05AX, N05BA, N05BB, N05BE, N05CC, N05CD, N05CF, N05CH, N05CM, N06AA, N06AB, N06AF, N06AG, N06AX, N06BA, N06BC, N06DA, N06DX, N07AA, N07AX, N07BA, N07BB, N07BC, N07CA, N07XX, P01AB, P01BA, P01BB, P01BC, P01BE, P01BF, P02CA, P02CF, P02CX, P03AC, R01A, R01AA, R01AC, R01AD, R01AX, R02AX, R03AC, R03AK, R03AL, R03BA, R03BB, R03CC, R03DA, R03DC, R03DX, R05CB, R05DA, R06AA, R06AD, |

| |
|---|
| R06AE, R06AX, R07AB, R07AX, S01*, S01AA, S01AD, S01AE, S01AX, S01BA, S01BC, S01CA, S01EA, S01EB, S01EC, S01ED, S01EE, S01FA, S01FB, S01GA, S01GX, S01HA, S01LA, S01XA, S02*, S02AA, S02BA, S02CA, S02D, S02DC, S03CA, V01AA, V03AB, V03AC, V03AE, V03AF, V03AX, V04CD, V04CH, V04CX, V06*, V06D, V07A, V07AB, V07AC, V07AY, V08AB, V08CA, V08DA, V09AX, A01, A01AA, A01AB, A01AC, A01AD, A02AA, A02AD, A02AH, A02BA, A02BB, A02BC, A02BX, A02X, A03AA, A03AB, A03BA, A03BB, A03FA, A04AA, A04AD, A05AA, A05BA, A06AA, A06AB, A06AC, A06AD, A06AG, A06AH, A06AX, A07AA, A07BA, A07CA, A07DA, A07EA, A07EC, A07FA, A09AA, A10AB, A10AC, A10AD, A10AE, A10BA, A10BB, A10BD, A10BH, A10BJ, A10BK, A10BX, A11AA, A11CC, A11DA, A11E, A11EA, A11EB, A11GA, A11HA, A12AA, A12AX, A12BA, A12CA, A12CB, A12CC, A12CE, A12CX, A16AB, B01, B01AA, B01AB, B01AC, B01AD, B01AE, B01AF, B01AX, B02AA, B02BA, B02BB, B02BC, B02BD, B03A, B03AA, B03AB, B03AC, B03AE, B03BA, B03BB, B03XA, B05, B05AA, B05BA, B05BB, B05BC, B05CX, B05DA, B05DB, B05XA, B05XC, C01AA, C01BC, C01BD, C01CA, C01CE, C01CX, C01DA, C01DX, C01EB, C02AB, C02AC, C02CA, C02DB, C02DC, C02DD, C02KX, C03AA, C03AB, C03BA, C03CA, C03DA, C03EA, C03XA, C05AA, C05BA, C05BB, C05CA, C07AA, C07AB, C07AG, C08CA, C08DA, C08DB, C09AA, C09BA, C09CA, C09DA, C09DX, C09XA, C10AA, C10AB, C10AC, C10AD, C10AX, D01AC, D01AE, D01BA, D02AB, D02AC, D02AX, D05AA, D05AX, D05BA, D05BB, D06AA, D06AX, D06BA, D06BB, D06BX, D07AA, D07AB, D07AC, D07AD, D07BC, D07CA, D07CC, D07XC, D08AJ, D10AD, D10AE, D10AF, D10AX, D11AH, D11AX, G01AF, G02AB, G02AD, G02BA, G02CB, G02CX, G03AA, G03AB, G03AC, G03BA, G03CA, G03CX, G03DA, G03FA, G03HA, G03XB, G03XC, G04BD, G04BE, G04CA, G04CB, H01AA, H01AC, H01BA, H01BB, H01CB, H02AA, H02AB, H03AA, H03BA, H03BB, H04AA, H05AA, H05BA, H05BX, J01AA, J01CA, J01CE, J01CF, J01CR, J01DB, J01DC, J01DD, J01DH, J01EA, J01EB, J01EE, J01FA, J01FF, J01GB, J01MA, J01XA, J01XB, J01XC, J01XD, J01XE, J01XX, J02AA, J02AC, J02AX, J04AB, J04AC, J04AK, J04AM, J05AB, J05AF, J05AH, J05AJ, J05AR, J05AX, J06BA, J06BB, J07AG, J07AJ, J07AL, J07AM, J07BB, J07BC, J07BF, L, L01, L01AA, L01AC, L01AD, L01BA, L01BB, L01BC, L01CA, L01CB, L01CD, L01CE, L01CX, L01DB, L01DC, L01EB, L01EJ, L01EL, L01EX, L01XA, L01XB, L01XC, L01XD, L01XE, L01XF, L01XG, L01XX, L01XY, L02AE, L02BA, L02BB, L02BG, L02BX, L03AA, L03AB, L03AX, L04AA, L04AB, L04AC, L04AD, L04AX, M01AA, M01AB, M01AE, M01AH, M01AX, M02AA, M03AB, M03AC, M03AX, M03BB, M03BX, M03CA, M04AA, M04AB, M04AC, M05BA, M05BX, N01AF, N01AH, N01AX, N01BB, N02AA, N02AB, N02AE, N02AG, N02AJ, N02AX, N02B, N02BA, N02BE, N02BG, N02CC, N02CD, N02CX, N03AA, N03AB, N03AE, N03AF, N03AG, N03AX, N04AA, N04AB, N04BA, N04BB, N04BC, N04BD, N04BX, N05AA, N05AB, N05AD, N05AE, N05AF, N05AG, N05AH, N05AL, N05AN, N05AX, N05BA, N05BB, N05BE, N05CC, N05CD, N05CF, N05CH, N05CM, N06AA, N06AB, N06AG, N06AX, N06BA, N06BC, N06DA, N06DX, N07AA, N07AX, N07BA, N07BB, N07BC, N07CA, N07XX, P01AB, P01BA, P01BC, R01A, R01AA, R01AC, R01AD, R01AX, R03AC, R03AK, R03AL, R03BA, R03BB, R03CC, R03DA, R03DC, R03DX, R05CB, R05DA, R06AA, R06AD, R06AE, R06AX, R07AB, R07AX, S01, S01AA, S01AX, S01BA, S01BC, S01CA, S01EA, S01EB, S01EC, S01ED, S01EE, S01FA, S01FB, S01GA, S01GX, S01HA, S01LA, S01XA, S02AA, S02CA, S03CA, V03AB, V03AC, V03AE, V03AF, V04CH, V04CX, V06, V07A, V07AB, V07AC, V07AY, V08AB, V08CA, V08DA, V09AX |
| **Laboratory tests represented as acronyms and local variable names**<br>25-Hydroxy-Vitamin D2;P, 25-Hydroxy-Vitamin D3;P, 25-OH-vitamin D (D3+D2);P, 3,4-Methylendioxyamfetamin;U, 3-Hydroxybutyrat;P, AFP, AGASBASE, AGASCO2, AGASHCO2, AGASLAKTAT, AGASO2, AGASPH, AGASSAT, ALB, ALP, ALT, AMYL, ANA, AV peak gradient, Acetoacetat (semikvant);U, Albumin / Kreatinin-ratio;U, Albumin;Csv, Albumin;Plv, Albumin;U, Amfetamin+analog;U, Ammonium;P, Amylase, pancreastype;P, Anion gap (inkl. K+);P, Antithrombin (enz.);P, Antitrombin (enz.);P, Antitrombin (koag.);P, Antitrombin;P, Ao V max, Ao V2 VTI, Ao V2 max PG, Ao V2 max vel, Aspartattransaminase [ASAT];P, Aspergillus (galactomannan Ag);P, Autofortolkning, B2M, BAC-test;B, BAS, BCx, BF-test udløb;P, BIL, BLYM, BSA, BUN, Bacterium+fungus;B(kateter; hæmodial.), Bacterium, nitrit-prod. (semikvant);U, Basisk fosfatase;P, Benzodiazepiner;U, Benzoylecgonin;U, Blastceller (uspec.);B, Bloddyrkning (Bakterium+fungus);aB, Bloddyrkning (fungus);B(CVK), Bloddyrkning (fungus);aB, Blodtype (AB0; Rh D);Erc(B), Brain natriuretisk peptid [BNP];P, Buprenorphin;U, C3, CARDIOLIPINIGG, CARDIOLIPINIGM, CD3, CD4, CD8, CHD-4-IgG [Mi-2];P, CICLO, CK, CKMB, CMVIGG, CMVPCR, CO2 total;P(vB), CORONAVIRUS SARS-COV-2 TOTAL IG, CRE, CRP, Calcium (albuminkorrigeret);P, Calcium-ion (frit)(pH=7,40; kont.renal erstat.terapi);P(vB;efter filter), Calcium-ion frit (pH = 7,40);P(vB;efter filter), Calcium-ion frit;P, Calcium-ion frit;P(vB;efter filter), Calcium;P, Candida mannan (Ag);P, Candida mannan-Ab;P, Candida-relateret egenskab gruppe;P, Cannabinol;U, Carbonmonoxidhæmoglobin;Hb(B), Centromer-IgG;P, Cerebrospinalvæske gruppe;Csv, Combat-COVID19-24T, Combat-COVID19-Start, Cystatin C;P, Cytomegalovirus-Ab;P, DDIM, DIFFBERE, DNA topoisomerase1-IgG [Scl70];P, DNAIGG, Deoxyhæmoglobin;Hb(tot.;aB), Digoxin;P, Direkte antiglobulin gruppe;Erc(B), Diverse analyse til KBA, EBNA, EBVPCR, EGFR, EOS, EVOL, EXOSC10-IgG [PM-Scl100];P, Eosinofilocytter;Csv, Erythrocyt-Ab gruppe;P, Erythrocytter (semikvant.);U, Erytroblaster;B, Erytrocytter;B, Erytrocytter;Csv, Erytrocytvol. rel. spredning;Erc(B), Erytrocytvolumen (middel) [MCV];B, Ethanol;P, FER, FIBR, Fibrillarin-IgG;P, Folat;P, Fosfat;P, GLUC, Gentamicin;P, Glomerulær basalmembran-IgG;P, Glukose (semikvant);U, Glukose;Csv, Glukose;P(aB), Glukose;P(kB), HAEM, HAPTO, HAVAB, HBVABC, HBVABS, HBVAGS, HBVIGM, HCG, HCVIGG, HDL, HGH CORONA - PAKKE, HIV 1+2 (Ag+Ab);P, Heparin lav molmasse (enz.);P, Histidin-tRNA-ligase (Jo1)-IgG;P, Hydrogencarbonat (akt.;Pt-tp);P(aB), Hydrogencarbonat;P(aB), Hydrogenkarbonat (aktuel);P(vB), Hæmoglobin (frit);P, Hæmoglobin (semikvant);U, Hæmoglobin A1c (IFCC);Hb(B), Hæmoglobin [MCHC];Erc(B), Hæmoglobinindhold [MCH];Erc(B), Hæmoglobinindhold;Rtcs(B), IGA, IGG, IGM, INR, IVS, Insulin;P(fPt), Interleukin-6;P, Jern;P, K+, KIA PROIL:TruCulture forskning;B, KIA profil: Multiplate, KIA profil: T-/B-/NK-lymfocytter;B, KOL, Kalium;Pt(U), Kalium;U, Kappa / Lambda-kæde (Ig) frit;P, Kappa-kæde (Ig) frit;P, Karbamid;Pt(U), Karbamid;U, |

| |
|---|
| Kerneholdige celler (uspec.);Csv, Kerneholdige celler;Csv, Kerneholdige celler;Plv, Keton;B, Klarhed før centrifugering;Csv, Klorid;P, Koag. TF-induceret tid;B, Koag. heparin-uafh tid;B, Koag. overflade-induceret (APTT)(BFH);P, Koag. overflade-induceret [APTT];P, Koag. overflade-induceret tid;B, Koagel-forskydningsstyrke;B, Koageldannelse, TF-induc.;B, Koagellyse, TF-induc.;B, Koagelstyrke, TF-induc.;B, Koagelstyrke, trombocyt-uafh;B, Koagulationsfaktor II+VII+X;P, Kolesterol LDL (beregnet);P, Kolesterol VLDL;P, Kolesterol non-HDL;P, Kortisol;P, Kreatinin-clearance;Nyre, Kreatinin;Pt(U), Kreatinin;U, LAC, LDH, LDL, LEUK, LV Mass Index, LV V1 max PG, LV mean PG, LVIDD, LVIDD index, LVIDS, LVOT diam, LVOT peak VTI, LYM, Laktat;Csv, Laktat;P, Laktat;P(vB), Laktatdehydrogenase;Plv, Lambda-kæde (Ig) frit;P, Leukoblaster;B, Leukocytter (mononukl.);Csv, Leukocytter (mononukl.);Plv, Leukocytter (polynukl.);Csv, Leukocytter (polynukl.);Plv, Leukocytter (semikvant);U, Leukocytter (uspec.);B, Leukocyttype gruppe;B, Levetiracetam;P, Lipase;P, Lithium;P, Lymfocytter+plasmaceller+blaster;B, Lymfocytter;Csv, M-komponent;P, MON, MV E/A, MV dec time, Magnesium;P, Major centromere B-IgG;P, Makrofager+monocytter;Csv, Markørundersøgelse, immundefekt projekt, Metamyelo.+Myelo.+Promyelocytter;B, Metamyelo.+myelo.+promyelocytter;B, Metamyelocytter;B, Methadon;U, Methæmoglobin;Hb(B), Mitose spindel-IgG;P, Monocytter+blaster;B, Morphin+analog;U, Multiplate-ADP;Trc(B), Multiplate-ASPI;Trc(B), Multiplate-TRAP(max);TRC(B), Myelocytter;B, Myoglobin;P, NA+, NEU, NK, Natrium;Pt(U), Natrium;U, Neutrofilocytter;Csv, Neutrophilocytter;B, Nitrit (semikvant);U, Nitrit;U, Nøgne kerner;B, O2 sat.;Hb(B), O2 sat.;Hb(aB;pulm.), O2 sat.;Hb(cvB), O2 sat.;Hb(kB), O2 sat.;Hb(vB), O2 sat;Hb(vB;pulm.), O2-flow;Pt, ORDERGROUP_EBV, Osmolalitet;P, Osmolalitet;U, Oxyhæmoglobin;Hb(aB;pulm.), Oxyhæmoglobin;Hb(tot.; aB), Oxyhæmoglobin;Hb(tot.; aB), Oxyhæmoglobin;Hb(tot.;aB), Oxyhæmoglobin;Hb(tot.;aB), Oxyhæmoglobin;Hb(tot.; vB), Oxyhæmoglobin;Hb(tot.; vB), Oxyhæmoglobin;Hb(tot.;vB), Oxyhæmoglobin;Hb(tot.;vB), Oxyhæmoglobin;Hb(tot.;cvB), P akse, P taks varighed, PR interval, PROCAL, PV peak gradient, PW, Paracetamol;P, Parathyrin [PTH];P, Plasmocytter;B, Pleuravæske gruppe;Plv, Pro-brain natriuretisk pept. [proBNP];P, Proinsulin C-peptid;P, Proinsulin C-peptid;P(fPt), Prolifererende nucleus-IgG;P, Promyelocytter;B, Prostataspecifikt antigen;P, Protein (semikvant);U, Protein;Csv, Protein;P, Protein;Plv, Protein;Pt(U), Protein;U, Proteinase 3-IgG [PR3];P, Prøvemateriale, QRS interval, QT interval, QTc (Bassett's formel), QTc (Fridericia's formel), R akse, RNA pol III RPC1-IgG;P, RR interval, RSJ SÆRAFTALE 00461, Reticulocytter gruppe;B, Reticulocytter;B, Reticulocytter;Erc(B), Ribosomal protein-IgG [Rib P];P, Rotem, Sedimentationsreaktion;B, Sjøgren syndrom [SSA]-IgG;P, Sjøgren syndrom [SSB]-IgG;P, Smiths-IgG;P, Smudge celler;Csv, Store ufarvede celler;B, Syrebasestatus gruppe;Pt, Syrebasestatus gruppe;Pt(aB), Syrebasestatus gruppe;Pt(vB), T akse, T-lymphocyt (helper/cytotox);Lymc(B), TEG, TEG FF-MA, TEG FFH-MA, TEG-Angle, TEG-LY30, TEG-MA, TEG-R, TEG-hep-Angle;B, TEG-hep-LY30;B, TEG-hep-MA;B, TEG-hep-R, TEG-heparinase, TR Max Vel, TR max PG, Tacrolimus;B, Thyrotropin [TSH]-reflextest;P, Thyrotropin [TSH];P, Thyroxin [T4];P, Thyroxin frit [T4];P, Transferrin-mætning;P, Transferrin;P, Triglycerid;P, Triiodthyronin [T3];P, Triiodthyronin frit [T3];P, Trombocytter;B, Troponin I;P, Troponin T;P, Troponin;P, U-PAR;P, U1 snRNP (70 kDa+A+C)-IgG;P, Urat;P, Urinopsamlingstid;Pt, Urinundersøgelse stix gruppe;U, Vancomycin;P, Ventrikelfrekvens, Vitamin B12;P, Volumen;Pt(U), Voriconazol;P, Zink;P, eAG, gamma-Glutamyltransferase [GGT];P, pCO2;P, pCO2;P(aB;pulm.), pCO2;P(cvB), pCO2;P(kB), pCO2;P(vB), pCO2;P(vB;pulm.), pH;P, pH;P(aB;pulm.), pH;P(cvB), pH;P(kB), pH;P(vB), pH;U, pO2 (halvmætn.);Hb(B), pO2;P, pO2;P(aB;pulm.), pO2;P(cvB), pO2;P(kB), pO2;P(vB), pO2;P(vB;pulm.), suPAR;P |
| **Vital parameters**<br>Blood pressure (diastolic), blood pressure (systolic), Glascow coma scale (GSC), Early Warning Score total, oxygen supply (L/min.), Pulse, Respiratory frequency (per minute), Oxygen Saturation, Temperature, Body Mass Index. |
| **Demographics**<br>Sex, Age, Pandemic wave, Pandemic week |
| **Hospitalizations**<br>Admitted at the time of first positive test, Previous admissions in the last 3 years, Cumulative days in hospital in the last 3 years |
| **Summary features**<br>Number of diagnoses, Number of ordered medicines, Number of administered medicines |

**Supplementary Table 2. Full set of features before feature selection**

Feature names and codes for the 2723 initial features that were used for feature selection. Values of the features were encoded using different time windows prior to a first positive test (FPT) and summary metrics according to the data type as detailed in text and figure 1. For vital parameters and laboratory tests, the last value within one month before FPT was used. For the case of diagnoses and medications, the total count of assigned codes within the last 3 and 1 year(s) before FPT, respectively, was encoded. Hospitalizations within the last 3 years before FPT were considered. Summary features were generated by counting the total number of diagnoses and medicine codes assigned to a patient.

| *Parameter* | *Value* |
|---|---|
| *boosting_type* | gbdt |
| *colsample_bytree* | 1 |
| *importance_type* | split |
| *learning_rate* | 0.05 |
| *max_depth* | -1 |
| *min_child_samples* | 20 |
| *min_child_weight* | 0.001 |
| *min_split_gain* | 0 |
| *n_estimators* | 100 |
| *n_jobs* | -1 |
| *num_leaves* | 31 |
| *objective* | binary |
| *reg_alpha* | 0 |
| *reg_lambda* | 0 |
| *silent* | TRUE |
| *subsample* | 0.7 |
| *subsample_for_bin* | 200000 |
| *subsample_freq* | 0 |
| *num_iterations* | 50 |
| *scale_pos_weight* | 100 |
| *metric* | auc |
| *seed* | 1234 |

**Supplementary Table 3. Hyperparameters for LightGBM classifiers.**
